# Maternal underweight and obesity are associated with placental pathologies in human pregnancy

**DOI:** 10.1101/2021.06.01.21258127

**Authors:** Hailey Scott, David Grynspan, Laura N Anderson, Kristin L Connor

**Author notes:** Correspondence: Dr. Kristin Connor, Department of Health Sciences, Carleton University, Ottawa, Ontario, Canada, K1S5B6 E.

## Abstract

**Introduction:** Maternal underweight and obesity are prevalent conditions, associated with chronic, low-grade inflammation, poor fetal development, and long-term adverse outcomes for the child. The placenta senses and adapts to the pregnancy environment in an effort to support optimal fetal development. However, the mechanisms driving these adaptations, and the resulting placental phenotypes, are poorly understood. We hypothesised that maternal underweight and obesity would be associated with increased prevalence of placental pathologies in term and preterm pregnancies.

**Methods:** Data from 12,154 pregnancies were obtained from the Collaborative Perinatal Project, a prospective cohort study conducted from 1959 to 1974. Macro and microscopic placental pathologies were analysed across maternal prepregnancy body mass index (BMI) to assess differences in the presence of pathologies among underweight, overweight, and obese BMI groups compared to normal weight reference BMI at term and preterm. Placental pathologies were also assessed across fetal sex.

**Results:** Pregnancies complicated by obesity had placentae with increased fetal inflammation at preterm, and increased inflammation of maternal gestational tissues at term. In term pregnancies, increasing maternal BMI associated with increased maternal vascular malperfusion (MVM), odds of an appropriately mature placenta for gestational age, and placental weight, and decreased placental efficiency. Male placentae, independent of maternal BMI, had increased inflammation, MVM, and placental efficiency than female placentae, particularly at term.

**Discussion:** Maternal underweight and obesity are not inert conditions for the placenta, and the histomorphological changes driven by suboptimal maternal BMI may serve as indicators of adversities experienced *in utero* and potential predictors of future health trajectories.

## Introduction

Maternal underweight and obesity are global health burdens; maternal underweight remains a persisting problem, and the prevalence of obesity in pregnancy continues to rise^1-3^. Both conditions have been associated with higher levels of inflammation in the mother, which favour increased inflammation in the placenta^4^, suboptimal nutrient availability to the fetus^4^, and adverse pregnancy and offspring outcomes^5,6^. Yet, the mechanisms that drive these outcomes remain poorly understood.

The placenta is a critical regulator of the fetal environment and can adapt to mitigate harmful exposures, or maladapt to permit their adverse effects^7^. For example, inflammation can reduce placental area^8^ and impair spiral artery remodelling^9^, which can affect nutrient and gas exchange^8-11^ with potential consequences for the developing offspring^12-14^. Additionally, there is some evidence that maternal underweight and obesity may be unfavourable for placental development. Maternal obesity has been associated with chronic villitis^15,16^, both delayed and accelerated placental villous maturation^16^, and increased atheromas and villous infarcts^16,17^, pathologies which have been linked to placental insufficiency^18^, fetal growth restriction, and neurodevelopmental impairment^16^. The effects of maternal undernutrition or low BMI on placental pathologies are less well documented^19^. Animal models of undernutrition have shown evidence of abnormal placental vasculature and decreased fetal blood space, labyrinth, and junctional zone area^19^, which may impair placental transfer^20^. The pro-inflammatory state induced by maternal underweight and obesity may also directly influence placental function, for example through altered expression of placental nutrient transporters^21-23^. Thus, to regulate the fetal environment, the placenta senses and integrates signals from the maternal environment, which may impact placental pathology. One mechanism through which the placenta may sense changes in maternal pathophysiology associated with suboptimal maternal BMI is mechanistic/mammalian target of rapamycin (mTOR), to regulate nutrient transport and subsequently, fetal growth^24,25^. Maternal obesity has been shown to activate placental mTOR, associated with increased fetal growth^26^, while conversely, intrauterine growth restriction, linked to maternal underweight^27^, has been associated with reduced placental mTOR signalling activity^28^. This suggests that the placenta integrates multiple signals, including those indicative of maternal supply and fetal demand, to regulate nutrient transfer and fetal growth^29^. Further, suboptimal maternal BMI is associated with altered metabolic status, and maternal obesity associates with dysregulated levels of metabolic hormones, including leptin and insulin, that are known to be upstream regulators of placental mTOR, and can influence placental nutrient transport^26^. The low-grade, pro-inflammatory state associated with maternal obesity may also alter placental outcomes, through increased placental levels of pro-inflammatory cytokines^30-32^, placental immune cells^31^, and activation of innate immune signalling pathways^32-34^, potentially mediating inflammatory signals between the mother, placenta, and fetus in pregnancies complicated by suboptimal maternal BMI.

However, given the multifactorial pathophysiology associated with suboptimal maternal BMI, including inflammatory, metabolic, and vascular factors^35,36^, reflected pathological phenotypes may vary. Even in the absence of overt adverse outcomes, such as large or small for gestational age infants or preterm birth^37^, changes to placental morphology and function (even subtle) may still have lasting effects on offspring health trajectories^38,39^. Therefore, it is important to understand even subtle placental phenotypes in these common conditions, as placental structural and functional adaptations may serve as a record of adversities experienced *in utero*, and could help reveal the mechanisms through which these adversities affect the developing offspring. However, the placental morphological and histopathologic changes induced by maternal underweight and obesity, in the absence of other major comorbidities or adverse perinatal events, are poorly characterized.

We hypothesised that, compared to women of normal weight, maternal underweight and obesity prepregnancy would be associated with increased prevalence of placental pathologies in term and preterm pregnancies, and that there would be differences in pathology prevalence based on placental sex. Using data from the Collaborative Perinatal Project (CPP), our primary objective was to determine whether placental pathologies were more prevalent in term and preterm pregnancies complicated by suboptimal maternal prepregnancy BMI. Our findings help to quantify placental pathologies in common pregnancy conditions, and uncover the mechanisms linking poor maternal metabolic health with suboptimal fetal growth and development.

## Methods

### Study design and population

Secondary data analysis was conducted using data from the CPP, a prospective cohort study designed to identify relationships between pregnancy and perinatal risk factors and child outcomes (https://catalog.archives.gov/id/606622). The CPP was conducted from 1959 to 1974 at 12 hospitals across the United States, and collected pregnancy data through the prenatal period and delivery, and child outcomes for approximately 58,000 pregnancies^40^.

The primary exposure of interest was maternal prepregnancy BMI, specifically maternal underweight, overweight, and obesity, compared to normal weight as the reference category. BMI was classified according to the World Health Organization and American College of Obstetricians and Gynecologists guidelines, where maternal BMI is categorized as underweight (<18.5), normal weight (18.5–24.9), overweight (25–29.9), or obese (≥30). To evaluate the associations between maternal BMI and placental pathologies, the study sample was restricted to pregnancies with placental pathology data available, and maternal height and prepregnancy weight available to calculate maternal prepregnancy BMI. We included only singleton births from first pregnancies (parity and gravidity of zero), where fetal sex was documented as male or female. Gestational age below 24 weeks or above 43 weeks were excluded based on the limit of viability^41^, and due to morphological changes to the placenta such as those induced by cellular senescence^42^, and increased risk for fetal complications in post-term pregnancies^43^. The CPP calculated gestational age based on the last menstrual period to the nearest week. These selection criteria resulted in a sample of 12,154 pregnancies (Supplementary Figure 1).

### Placental pathologies

The primary outcomes were macroscopic and microscopic placental pathologies (Figure 1). Macroscopic data included placental weight, largest and smallest diameter, thickness, and placental shape. Infant birthweight to placental weight/largest diameter/smallest diameter/thickness ratios were calculated as potential predictors of placental efficiency^38,44^. The top and bottom 0.5% of raw infant and placental anthropometry data (birthweight, placental weight, and placental dimensions) were excluded to remove biologically implausible data. The umbilical cord was assessed for cord edema and number of vessels, given the increased incidence of a single artery cord with gestational diabetes mellitus (GDM)^45^, a covariate of interest for pregnancies with obesity. Thrombosed fetal vessels and cut surface infarcts were assessed, which may be associated with impaired placental perfusion^46^. For microscopic variables: decidual vessel fibrinoids and atheroma were included, features of malperfusion^47^. Neutrophilic infiltration of the umbilical vein, umbilical artery, cord substance, chorion and amnion membranes, and chorion and amnion of the (fetal side) placental surface were also obtained, which may be indicative of ascending maternal infection^48^. Cut surface calcification was included as an indicator for placental maturation^49^. Syncytium-nuclear clumping, or syncytial knots, and stromal fibrosis are signs of accelerated villous maturation and were thus included^50^. Prescence of Langhans’ layer, Hofbauer cells, and pathological edema were included as indicators of placental immaturity^51,52^. A variable provided for the apparent maturity of the placenta was also included as a marker for appropriate placental development for gestational age. For multivariable analyses, categorical placental pathology variables were collapsed into binary categories (Supplementary Table 1).

**Figure 1.**
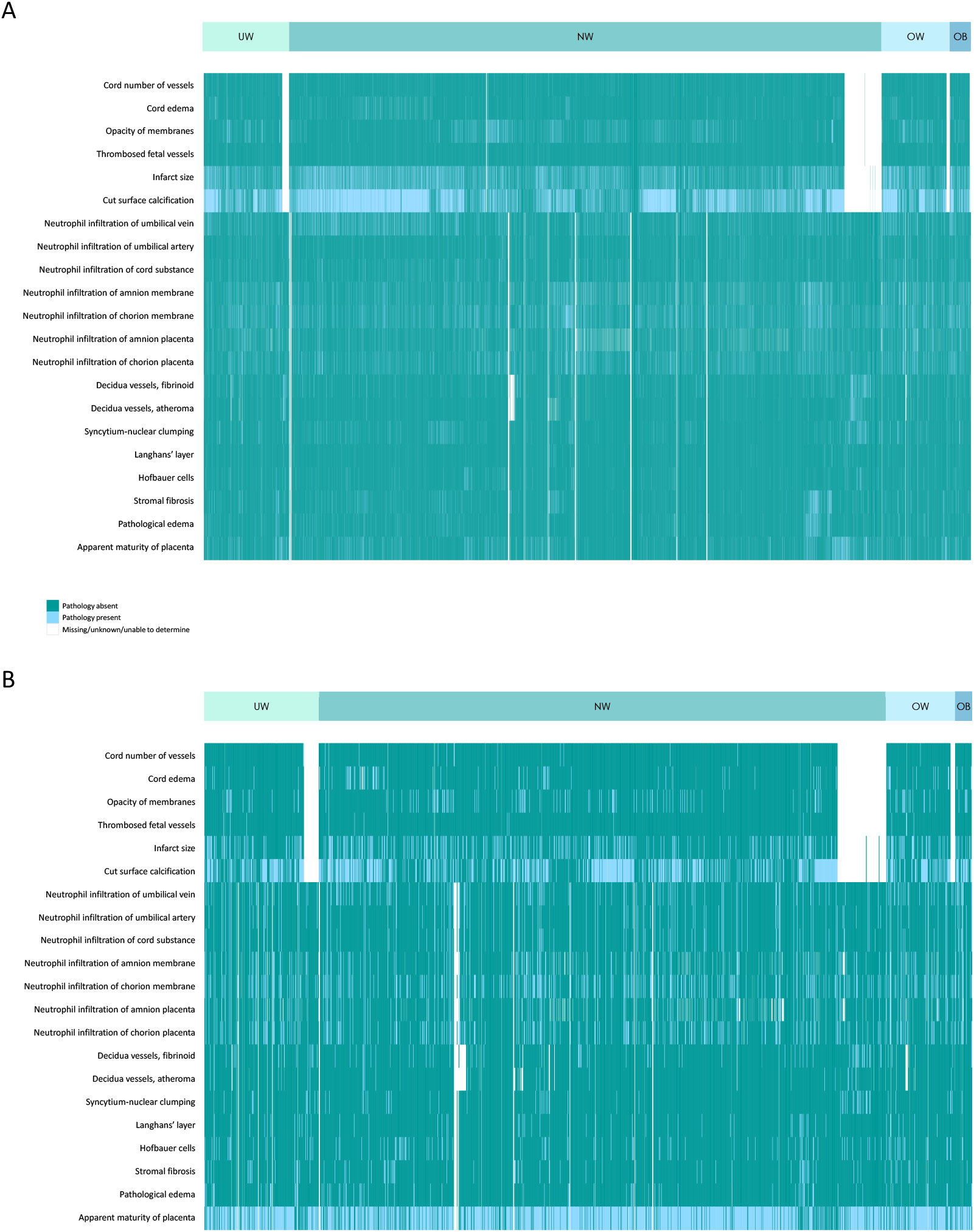
Prevalence of placental pathologies in term (A) and preterm (B) pregnancies. The presence or absence of categorical placental pathologies across maternal prepregnancy body mass index (BMI) groups, where mothers were categorized as underweight (UW), normal weight (NW), overweight (OW) or having obesity (OB), in term (n=10,415; panel A) and preterm (n=1,739; panel B) pregnancies.

We derived additional summary scores for placental inflammation, maternal vascular malperfusion (MVM), and placental immaturity. A maternal inflammation summary score was derived from the following variables: opacity of membranes and neutrophil infiltration of the amnion and chorion membranes and of the placental surface. Where data were available for all constituent variables, the individual variable scores were summed to create a composite score ranging from zero to fourteen, where a higher score represents increased levels of inflammation. Similarly, a fetal inflammation summary score was derived from neutrophilic infiltration of the umbilical vein, artery, and cord substance to create a composite score from zero to nine. A summary score considering features of MVM was derived from: presence of infarcts and syncytium-nuclear clumping to create a score ranging from zero to two, where a higher score indicates increased MVM. A placental immaturity score was derived from the following variables: presence of Langhans’ layer, Hofbauer cells, absence of stromal fibrosis, and normal or less than normal syncytium-nuclear clumping, to create a score ranging from zero to four where a higher score indicates a more immature placental phenotype (Supplementary Table 2).

### Maternal demographics

Our primary exposure of interest, maternal prepregnancy BMI, was defined based on measured height and self-reported prepregnancy weight collected at study enrollment. In addition to prepregnancy BMI, maternal demographic data including age, race, education, marital status, income, socioeconomic index, housing density, smoking history, diabetes mellitus status, and gestational weight gain were obtained. The socioeconomic index is a composite numerical index derived from scores for education (of the head of household/chief earner), occupation (of the head of household/chief earner), and family income, ranging from 0.0-9.5, where 9.5 represents the highest socioeconomic status^53^. Smoking history was provided as the number of cigarettes smoked per day at the time of the interview, from zero cigarettes (including non-smokers) to 60 cigarettes per day. Additional smoking categories included 61 or more cigarettes per day, regular smoker but less than one cigarette per day, and irregular smoker but less than four cigarettes per month. Smoking history was presented as non-smokers (including women currently smoking zero cigarettes per day), light smokers (less than one pack of 20 cigarettes per day), and heavy smokers (one or more packs of 20 cigarettes per day). Diabetes mellitus was presented as presence or absence, where presence included diabetes mellitus reported before pregnancy, during pregnancy, both before and during, during and postpartum, or before, during, and postpartum. Based on current recommendations from the Institute of Medicine (IOM) guidelines (2009), maternal weight gain was categorized as inadequate, adequate, or excessive for singleton pregnancies based on prepregnancy BMI, where the recommended weight gain ranges are 28–40 pounds, 25–35 pounds, 15–25 pounds or 11–20 pounds^54^ for mothers who are underweight, normal weight, overweight, or have obesity, respectively.

### Statistical analyses

#### Univariate analyses

Univariate analysis was conducted to evaluate differences in the prevalence of placental pathologies across maternal prepregnancy BMI groups. Given that the presence of some placental pathologies are dependent on gestational age^55^, we conducted all analyses stratified for term and preterm placentae. Differences between maternal BMI groups and placental measures were determined by Kruskal–Wallis test with Steel–Dwass *post hoc* for continuous nonparametric data, and Likelihood Ratio Chi Square test for categorical data. Placental pathologies were also assessed across fetal sex. Data are presented as median (interquartile range) and Wilcoxon test effect size (r) (95% confidence interval; CI) for non-parametric continuous data, and frequency (percentage) and Cramer’s V effect size (95% CI) or odds ratio (95% CI) (for binary fetal sex analyses) for categorical variables. Statistical significance was defined as p<0.05. Data were analysed using JMP statistical software (14.0), and Wilcoxon test effect size and Cramer’s V effect size were calculated in R (4.1.2).

#### Multivariable analyses

We performed multivariable logistic and linear regression to determine the relationships between maternal prepregnancy BMI and placental pathologies at term and preterm. Logistic regression models were used to determine the associations between maternal BMI (continuous) and binary placental pathologies. Categorical placental pathology variables were collapsed into binary categories to calculate odds ratios for the pathological phenotype. Any data coded as unknown, unable to determine, or missing were excluded from regression analyses. Data are presented as unadjusted (OR) or adjusted units odds ratio (aOR) (95% CI). Linear regression models were used to determine the associations between maternal BMI (continuous) and continuous placental pathology variables. Data are reported as adjusted beta coefficient (aβ) (95% CI). Covariates of interest were identified a priori and included fetal sex (male/female), maternal race (White, Black, and Other), maternal age (continuous), smoking history (non-smoker, light smoker, heavy smoker), maximum gestational weight gain (continuous), diabetes (yes/no), maternal education (continuous), and socioeconomic index (continuous). Two regression models were defined a priori: 1) An unadjusted model was first used to identify the associations between prepregnancy BMI alone (as a continuous variable) with placental pathologies and 2) an adjusted model adjusted for the covariates defined above.

## Results

### Maternal demographics differed across maternal BMI groups

Maximum gestational weight gain among preterm (Wilcoxon test effect size, r=-0.09 [-0.14 to -0.04]) and term (r=-0.05 [-0.07 to -0.03]) pregnancies was greatest in underweight and lowest in obese BMI groups (Supplementary Tables 3-4). However, according to IOM (2009) guidelines, weight gain in underweight pregnancies was still inadequate at preterm and term, while most pregnancies with obesity had either inadequate or excessive weight gain at preterm, and excessive weight gain at term (Supplementary Tables 3-4). Maternal age, race, education, marital status, income, socioeconomic index, and housing density also differed by BMI group (Supplementary Tables 3-4). There were no differences in median maternal prepregnancy BMI across study years (based on date of delivery, p=0.10). The median BMI from 1959 to 1966 ranged from 20.9 (in years 1960, 1962, 1963, 1966) to 21.3 (in 1959; Supplementary Figure 2). Socioeconomic index varied by study site (p<0.0001); across the 12 study sites, median socioeconomic index ranged from 3.0, relatively low on the socioeconomic index scale ranging from 0.0 to 9.5 (Virginia; Tennessee; Louisiana locations) to 8.3, a relatively high socioeconomic index score (Buffalo, New York location; Supplementary Table 5). There was no association between maternal BMI and socioeconomic index at preterm, but at term, socioeconomic index was highest among term normal weight pregnancies compared to all other BMI groups (p<0.0001, Supplementary Table 4).

### Maternal obesity associated with increased neutrophil infiltration of gestational tissues

Among preterm pregnancies, fetal inflammation, characterised by the composite fetal inflammation summary score, was greater in placentae from mothers with obesity compared to underweight and normal weight BMI groups (r=0.009 [-0.04 to 0.06], Table 1). There were no effects of maternal BMI group on neutrophil infiltration in the preterm membranes or placental surface. Neutrophil infiltration of the umbilical vein (aOR=1.06 [1.01–1.12]), umbilical cord substance (aOR=1.08 [1.02– 1.14]), and amnion membrane (OR=1.05 [1.01–1.10]), but not the umbilical artery, chorion membrane, or amnion and chorion of the placenta, were more likely with increasing maternal BMI (Table 2).

**Table 1.**
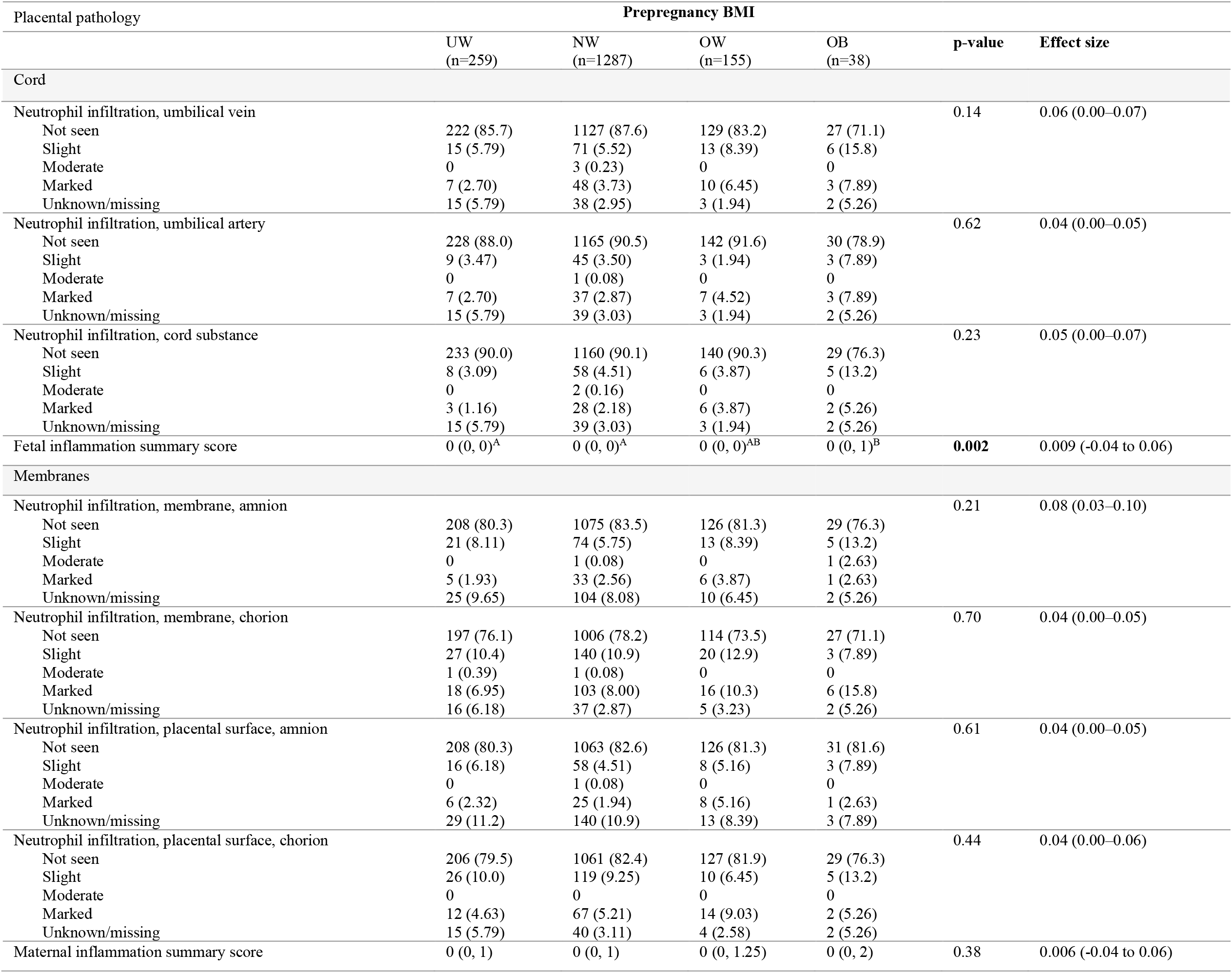
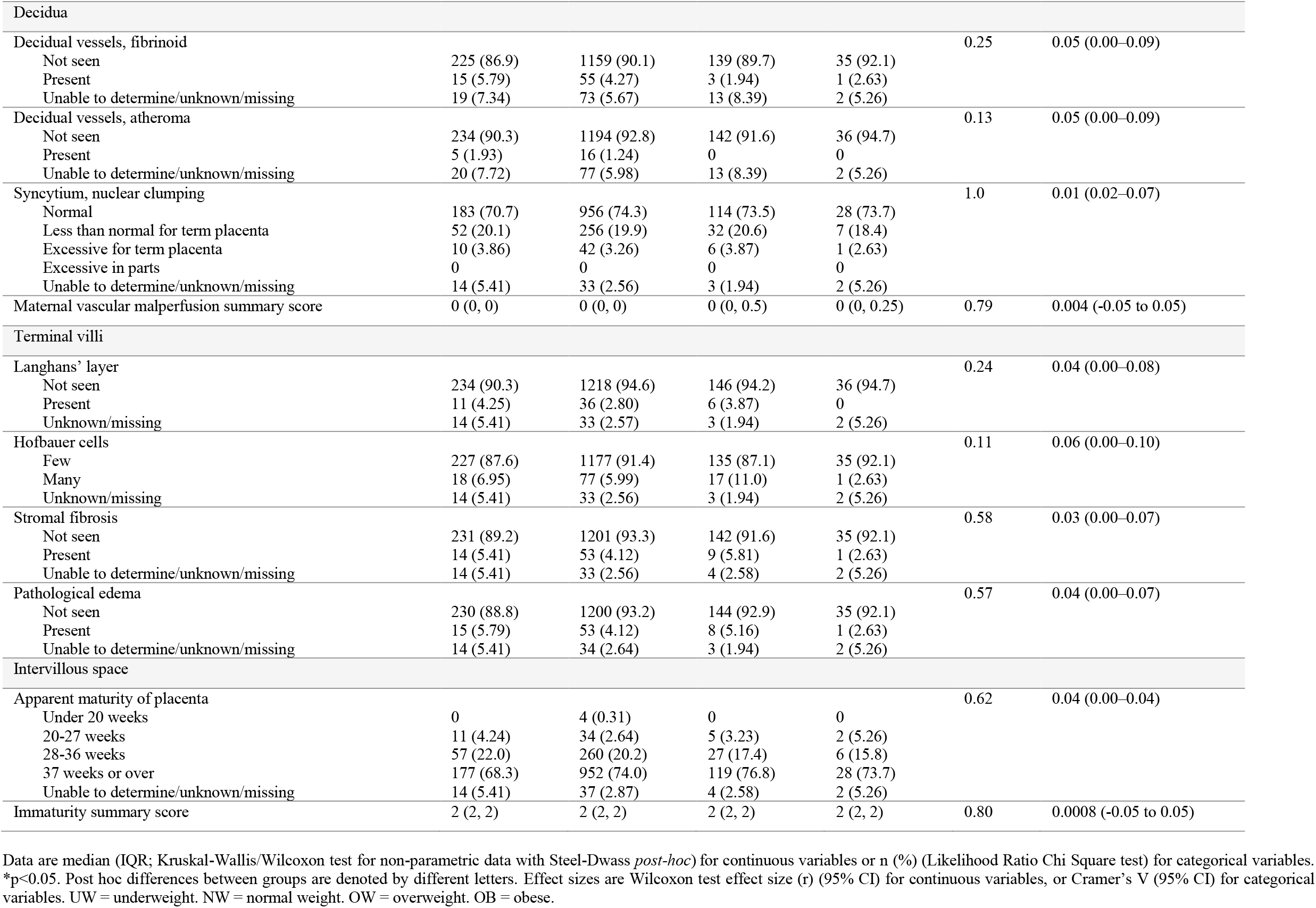
Associations between maternal prepregnancy BMI and microscopic placental pathologies in preterm pregnancies, N=1739.

**Table 2.**
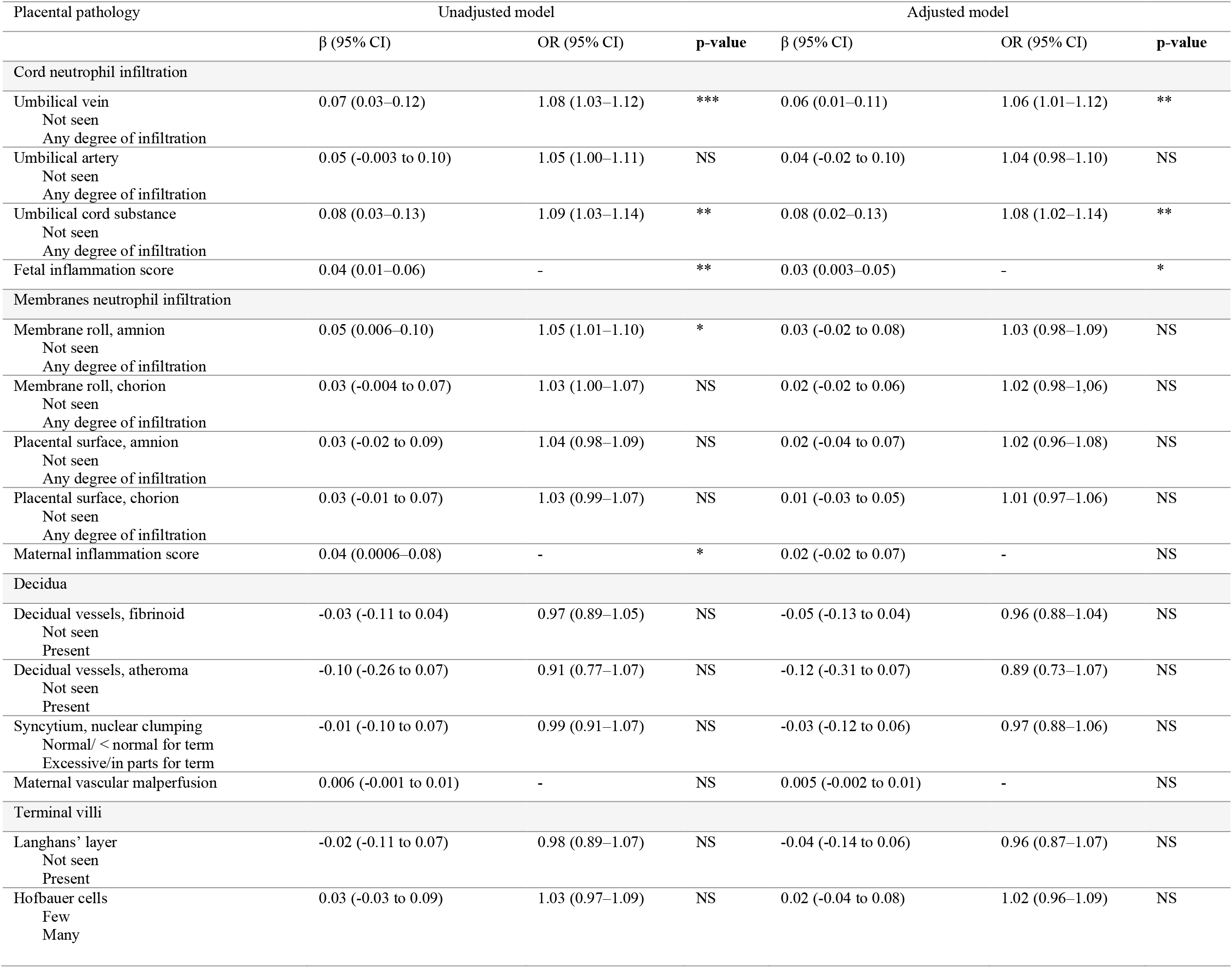
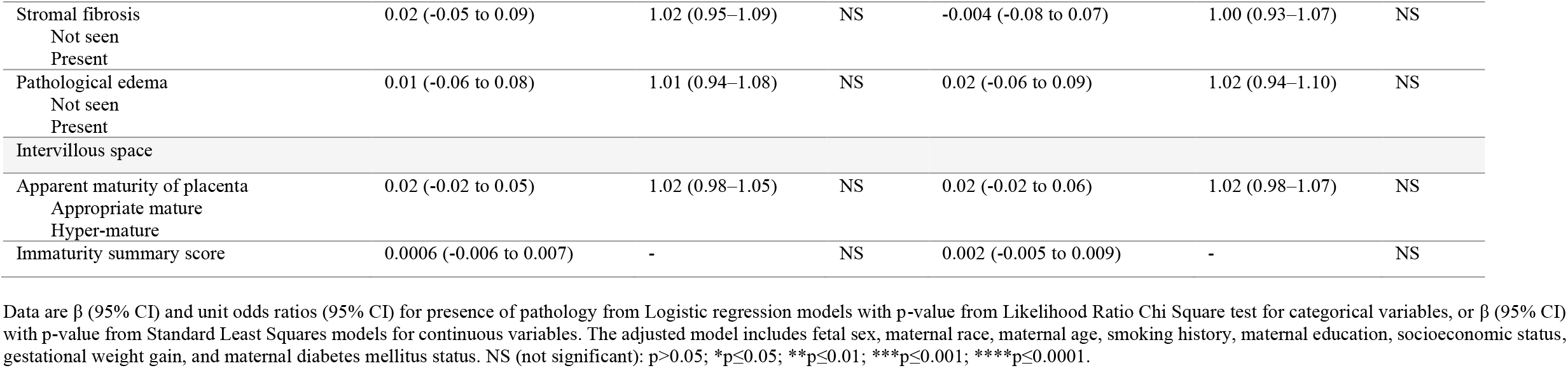
Multivariable analyses for associations between maternal prepregnancy BMI (continuous) and microscopic placental pathologies in preterm pregnancies, N=1739.

Among term pregnancies, pregnancies complicated by obesity had the greatest percentage of marked neutrophil infiltration of the umbilical vein and artery, amnion and chorion membranes, and amnion, but not chorion, of the placental surface (Table 3). The maternal inflammation summary score was greatest among pregnancies complicated by obesity, though there were no differences between BMI groups on post hoc analysis (r=0.008 [-0.01 to 0.03], Table 3). Further, neutrophilic infiltration of the umbilical vein (aOR=1.03 [1.01–1.05]), artery (aOR=1.04 [1.00–1.07]), and cord substance (aOR=1.04 [1.01–1.07]) increased with increasing BMI (Table 4). Neutrophil infiltration of the amnion membrane (aOR=1.06 [1.03–1.08]), chorion membrane (aOR=1.04 [1.02– 1.06]), amnion of the placental surface (aOR=1.04 [1.01–1.07]), and chorion of the placental surface (aOR=1.04 [1.02–1.06]) were also greater with increasing BMI (Table 4).

**Table 3.** Associations between maternal prepregnancy BMI and microscopic placental pathologies in term pregnancies, N=10,415.

| Placental pathology | Prepregnancy BMI |  |  |  | p-value | Effect size |
| --- | --- | --- | --- | --- | --- | --- |
|  | UW<br>(n=1201) | NW<br>(n=8022) | OW<br>(n=912) | OB<br>(n=280) |  |  |
| Cord |  |  |  |  |  |  |
| Neutrophil infiltration, umbilical vein |  |  |  |  | 0.001 | 0.03 (0.01–0.04) |
| Not seen | 1065 (88.7) | 6965 (86.8) | 787 (86.3) | 236 (84.3) |  |  |
| Slight | 84 (6.99) | 604 (7.53) | 51 (5.59) | 22 (7.86) |  |  |
| Moderate | 0 | 16 (0.20) | 0 | 0 |  |  |
| Marked | 24 (2.0) | 198 (2.47) | 35 (3.84) | 15 (5.36) |  |  |
| Unknown/missing | 28 (2.33) | 239 (2.98) | 39 (4.28) | 7 (2.50) |  |  |
| Neutrophil infiltration, umbilical artery |  |  |  |  | 0.05 | 0.03 (0.006–0.03) |
| Not seen | 1141 (95.0) | 7563 (94.3) | 840 (92.1) | 259 (92.5) |  |  |
| Slight | 20 (1.67) | 142 (1.77) | 16 (1.75) | 5 (1.79) |  |  |
| Moderate | 0 | 3 (0.04) | 0 | 0 |  |  |
| Marked | 12 (1.0) | 75 (0.93) | 18 (1.97) | 9 (3.21) |  |  |
| Unknown/missing | 28 (2.33) | 239 (2.98) | 38 (4.17) | 7 (2.50) |  |  |
| Neutrophil infiltration, umbilical cord substance |  |  |  |  | 0.09 | 0.02 (0.00–0.03) |
| Not seen | 1130 (94.1) | 7442 (92.8) | 825 (90.5) | 252 (90.0) |  |  |
| Slight | 34 (2.83) | 265 (3.30) | 36 (3.95) | 13 (4.64) |  |  |
| Moderate | 0 | 5 (0.06) | 1 (0.11) | 0 |  |  |
| Marked | 9 (0.75) | 71 (0.89) | 12 (1.32) | 8 (2.86) |  |  |
| Unknown/missing | 28 (2.33) | 239 (2.98) | 38 (4.12) | 7 (2.50) |  |  |
| Fetal inflammation summary score | 0 (0, 0) | 0 (0, 0) | 0 (0, 0) | 0 (0, 0) | 0.10 | 0.01 (-0.008 to 0.03) |
| Membranes |  |  |  |  |  |  |
| Neutrophil infiltration, membrane roll, amnion |  |  |  |  | 0.002 | 0.03 (0.01–0.04) |
| Not seen | 1028 (85.6) | 6736 (84.0) | 732 (80.3) | 223 (79.6) |  |  |
| Slight | 62 (5.16) | 434 (5.41) | 71 (7.79) | 25 (8.93) |  |  |
| Moderate | 0 | 5 (0.06) | 0 | 1 (0.36) |  |  |
| Marked | 16 (1.33) | 158 (1.97) | 24 (2.63) | 9 (3.21) |  |  |
| Unknown/missing | 95 (7.91) | 689 (8.59) | 85 (9.32) | 22 (7.86) |  |  |
| Neutrophil infiltration, membrane roll, chorion |  |  |  |  | 0.006 | 0.03 (0.009–0.04) |
| Not seen | 1032 (85.9) | 6718 (83.7) | 742 (81.36) | 216 (77.1) |  |  |
| Slight | 97 (8.08) | 763 (9.51) | 87 (9.54) | 40 (14.3) |  |  |
| Moderate | 2 (0.17) | 9 (0.11) | 0 | 0 |  |  |
| Marked | 39 (3.25) | 288 (3.59) | 45 (4.93) | 18 (6.43) |  |  |
| Unknown/missing | 31 (2.58) | 244 (3.04) | 38 (4.17) | 6 (2.14) |  |  |
| Neutrophil infiltration, placental surface, amnion |  |  |  |  | 0.06 | 0.03 (0.005–0.03) |
| Not seen | 1011 (84.2) | 6691 (83.4) | 740 (81.1) | 236 (84.3) |  |  |
| Slight | 44 (3.66) | 290 (3.62) | 45 (4.93) | 13 (4.64) |  |  |
| Moderate | 1 (0.08) | 2 (0.02) | 1 (0.11) | 1 (0.36) |  |  |
| Marked | 9 (0.75) | 96 (1.20) | 17 (1.86) | 7 (2.50) |  |  |
| Unknown/missing | 136 (11.32) | 943 (11.8) | 109 (12.0) | 23 (8.21) |  |  |
| Neutrophil infiltration, placental surface, chorion |  |  |  |  | 0.08 | 0.02 (0.00–0.03) |
| Not seen | 1070 (89.1) | 6983 (87.0) | 766 (84.0) | 239 (85.4) |  |  |
| Slight | 85 (7.08) | 624 (7.78) | 80 (8.77) | 21 (7.50) |  |  |
| Moderate | 1 (0.08) | 6 (0.07) | 1 (0.11) | 1 (0.36) |  |  |
| Marked | 19 (1.58) | 186 (2.32) | 29 (3.18) | 13 (4.64) |  |  |
| Unknown/missing | 26 (2.16) | 223 (2.78) | 36 (3.95) | 6 (2.14) |  |  |
| Maternal inflammation summary score | 0 (0, 0)<sup>A</sup> | 0 (0, 0)<sup>A</sup> | 0 (0, 0)<sup>A</sup> | 0 (0, 1)<sup>A</sup> | 0.02 | 0.008 (-0.01 to 0.03) |
| Decidua |  |  |  |  |  |  |
| Decidual vessels, fibrinoid |  |  |  |  | 0.21 | 0.02 (0.00–0.04) |
| Not seen | 1106 (92.1) | 7417 (92.5) | 838 (91.9) | 258 (92.1) |  |  |
| Present | 44 (3.66) | 226 (2.82) | 20 (2.19) | 6 (2.14) |  |  |
| Unable to determine/ unknown/missing | 51 (4.25) | 379 (4.72) | 54 (5.92) | 16 (5.71) |  |  |
| Decidual vessels, atheroma |  |  |  |  | 0.25 | 0.02 (0.00–0.04) |
| Not seen | 1127 (93.8) | 7533 (93.9) | 844 (92.5) | 260 (92.9) |  |  |
| Present | 21 (1.75) | 83 (1.03) | 10 (1.10) | 3 (1.07) |  |  |
| Unable to determine/ unknown/missing | 53 (4.41) | 406 (5.06) | 58 (6.36) | 17 (6.07) |  |  |
| Syncytium, nuclear clumping |  |  |  |  | 0.59 | 0.02 (0.00–0.02) |
| Normal | 1046 (87.1) | 6973 (86.9) | 790 (86.6) | 251 (89.6) |  |  |
| Less than normal for term placenta | 97 (8.08) | 556 (6.93) | 60 (6.58) | 13 (4.64) |  |  |
| Excessive for term placenta | 29 (2.41) | 238 (2.97) | 21 (2.30) | 9 (3.21) |  |  |
| Excessive in parts | 5 (0.41) | 41 (0.51) | 5 (0.55) | 1 (0.36) |  |  |
| Unable to determine/unknown/missing | 24 (2.00) | 214 (2.67) | 36 (3.95) | 6 (2.14) |  |  |
| Maternal vascular malperfusion summary score | 0 (0, 1) | 0 (0, 1) | 0 (0, 1) | 0 (0, 1) | 0.11 | 0.02 (-0.004 to 0.04) |
| Terminal villi |  |  |  |  |  |  |
| Langhans' layer |  |  |  |  | 0.39 | 0.02 (0.00–0.03) |
| Not seen | 1173 (97.7) | 7765 (96.8) | 869 (95.3) | 271 (96.8) |  |  |
| Present | 4 (0.33) | 46 (0.57) | 7 (0.77) | 3 (1.07) |  |  |
| Unknown/missing | 24 (2.00) | 211 (2.63) | 36 (3.95) | 6 (2.14) |  |  |
| Hofbauer cells |  |  |  |  | 0.91 | 0.007 (0.00–0.02) |
| Few | 1155 (96.2) | 7672 (95.6) | 862 (94.5) | 269 (96.1) |  |  |
| Many | 22 (1.83) | 140 (1.75) | 13 (1.43) | 5 (1.79) |  |  |
| Unknown/missing | 24 (2.00) | 210 (2.62) | 37 (4.06) | 6 (2.14) |  |  |
| Stromal fibrosis |  |  |  |  | 0.25 | 0.02 (0.00–0.04) |
| Not seen | 1135 (94.5) | 7499 (93.5) | 851 (93.3) | 261 (93.2) |  |  |
| Present | 41 (3.41) | 313 (3.90) | 25 (2.74) | 13 (4.64) |  |  |
| Unable to determine/unknown/missing | 25 (2.08) | 210 (2.62) | 36 (3.95) | 6 (2.14) |  |  |
| Pathological edema |  |  |  |  | 0.12 | 0.02 (0.00–0.04) |
| Not seen | 1135 (94.5) | 7553 (94.2) | 847 (92.9) | 271 (96.8) |  |  |
| Present | 42 (3.50) | 258 (3.22) | 29 (3.18) | 3 (1.07) |  |  |
| Unable to determine/unknown/missing | 24 (2.00) | 211 (2.63) | 36 (3.95) | 6 (2.14) |  |  |
| Intervillous space |  |  |  |  |  |  |
| Apparent maturity of placenta |  |  |  |  | 0.72 | 0.01 (0.00–0.01) |
| Under 20 weeks | 1 (0.08) | 6 (0.07) | 0 | 0 |  |  |
| 20-27 weeks | 4 (0.33) | 19 (0.24) | 1 (0.11) | 0 |  |  |
| 28-36 weeks | 92 (7.66) | 543 (6.77) | 55 (6.03) | 19 (6.79) |  |  |
| 37 weeks or over | 1078 (89.8) | 7237 (90.2) | 820 (89.9) | 255 (91.1) |  |  |
| Unable to determine/unknown/missing | 26 (2.16) | 217 (2.71) | 36 (3.95) | 6 (2.14) |  |  |
| Immaturity summary score | 2 (2, 2) | 2 (2, 2) | 2 (2, 2) | 2 (2, 2) | 0.35 | -0.01 (-0.03 to 0.007) |

Data are median (IQR; Kruskal-Wallis/Wilcoxon test for non-parametric data with Steel-Dwass *post-hoc*) for continuous variables or n (%) (Likelihood Ratio Chi Square test) for categorical variables. \*p<0.05. Post hoc differences between groups are denoted by different letters. Effect sizes are Wilcoxon test effect size (r) (95% CI) for continuous variables, or Cramer's V (95% CI) for categorical variables. UW = underweight. NW = normal weight. OW = overweight. OB = obese.

**Table 4.** Multivariable analyses for associations between maternal prepregnancy BMI (continuous) and microscopic placental pathologies in term pregnancies, N=10,415.

| Placental pathology | Unadjusted model |  |  | Adjusted model |  |  |
| --- | --- | --- | --- | --- | --- | --- |
| | $\beta$ (95% CI) | OR (95% CI) | p-value | $\beta$ (95% CI) | OR (95% CI) | p-value |
| **Cord neutrophil infiltration** |  |  |  |  |  |  |
| Umbilical vein | 0.02 (0.004–0.04) | 1.02 (1.00–1.04) | * | 0.03 (0.007–0.05) | 1.03 (1.01–1.05) | ** |
| Not seen |  |  |  |  |  |  |
| Any degree of infiltration |  |  |  |  |  |  |
| Umbilical artery | 0.05 (0.02–0.08) | 1.05 (1.02–1.08) | ** | 0.04 (0.003–0.07) | 1.04 (1.00–1.07) | * |
| Not seen |  |  |  |  |  |  |
| Any degree of infiltration |  |  |  |  |  |  |
| Umbilical cord substance | 0.04 (0.02–0.07) | 1.04 (1.02–1.07) | *** | 0.04 (0.008–0.06) | 1.04 (1.01–1.07) | ** |
| Not seen |  |  |  |  |  |  |
| Any degree of infiltration |  |  |  |  |  |  |
| Fetal inflammation score | 0.01 (0.008–0.02) | - | **** | 0.01 (0.007–0.02) | - | **** |
| **Membranes and neutrophil infiltration** |  |  |  |  |  |  |
| Membrane roll, amnion | 0.06 (0.04–0.07) | 1.06 (1.04–1.08) | **** | 0.05 (0.03–0.08) | 1.06 (1.03–1.08) | **** |
| Not seen |  |  |  |  |  |  |
| Any degree of infiltration |  |  |  |  |  |  |
| Membrane roll, chorion | 0.05 (0.03–0.06) | 1.05 (1.03–1.07) | **** | 0.04 (0.02–0.06) | 1.04 (1.02–1.06) | **** |
| Not seen |  |  |  |  |  |  |
| Any degree of infiltration |  |  |  |  |  |  |
| Placental surface, amnion | 0.04 (0.01–0.06) | 1.04 (1.01–1.06) | ** | 0.04 (0.01–0.06) | 1.04 (1.01–1.07) | ** |
| Not seen |  |  |  |  |  |  |
| Any degree of infiltration |  |  |  |  |  |  |
| Placental surface, chorion | 0.04 (0.02–0.06) | 1.04 (1.02–1.06) | **** | 0.04 (0.02–0.06) | 1.04 (1.02–1.06) | *** |
| Not seen |  |  |  |  |  |  |
| Any degree of infiltration |  |  |  |  |  |  |
| Maternal inflammation score | 0.03 (0.02–0.04) | - | **** | 0.03 (0.02–0.04) | - | **** |
| **Decidua** |  |  |  |  |  |  |
| Decidual vessels, fibrinoid | -0.03 (-0.07 to 0.01) | 0.97 (0.94–1.01) | NS | -0.02 (-0.06 to 0.02) | 0.98 (0.94–1.02) | NS |
| Not seen |  |  |  |  |  |  |
| Present |  |  |  |  |  |  |
| Decidual vessels, atheroma | -0.05 (-0.12 to 0.01) | 0.95 (0.89–1.01) | NS | -0.06 (-0.13 to 0.004) | 0.94 (0.88–1.00) | NS |
| Not seen |  |  |  |  |  |  |
| Present |  |  |  |  |  |  |
| Syncytium, nuclear clumping | 0.002 (-0.03 to 0.03) | 1.00 (0.97–1.03) | NS | 0.001 (-0.03 to 0.04) | 1.00 (0.97–1.04) | NS |
| Normal/ < normal for term |  |  |  |  |  |  |
| Excessive/in parts for term |  |  |  |  |  |  |
| Maternal vascular malperfusion | 0.005 (0.002–0.008) | - | ** | 0.007 (0.004–0.01) | - | **** |
| **Terminal villi** |  |  |  |  |  |  |
| Langhans' layer | 0.007 (-0.07 to 0.08) | 1.01 (0.93–1.09) | NS | -0.008 (-0.09 to 0.07) | 0.99 (0.92–1.07) | NS |
| Not seen |  |  |  |  |  |  |
| Present |  |  |  |  |  |  |
| Hofbauer cells | -0.03 (-0.08 to 0.02) | 0.97 (0.92–1.02) | NS | -0.04 (-0.09 to 0.009) | 0.96 (0.91–1.01) | NS |
| Few |  |  |  |  |  |  |
| Many |  |  |  |  |  |  |
| Stromal fibrosis | -0.006 (-0.04 to 0.03) | 0.99 (0.96–1.03) | NS | 0.002 (-0.03 to 0.04) | 1.00 (0.97–1.04) | NS |
| Not seen |  |  |  |  |  |  |
| Present |  |  |  |  |  |  |
| Pathological edema | -0.03 (-0.07 to 0.004) | 0.97 (0.93–1.00) | NS | -0.01 (-0.05 to 0.03) | 0.99 (0.95–1.03) | NS |
| Not seen |  |  |  |  |  |  |
| Present |  |  |  |  |  |  |
| Intervillous space |  |  |  |  |  |  |
| Apparent maturity of placenta | -0.02 (-0.05 to 0.003) | 0.98 (0.96–1.00) | NS | -0.06 (-0.09 to -0.03) | 0.94 (0.92–0.97) | **** |
| Appropriate mature |  |  |  |  |  |  |
| Immature |  |  |  |  |  |  |
| Immaturity score | -0.0003 (-0.002 to 0.002) | - | NS | -0.0009 (-0.003 to 0.001) | - | NS |

Data are $\beta$ (95% CI) and unit odds ratios (95% CI) for presence of pathology from Logistic regression models with p-value from Likelihood Ratio Chi Square test for categorical variables, or $\beta$ (95% CI) with p-value from Standard Least Squares models for continuous variables. The adjusted model includes fetal sex, maternal race, maternal age, smoking history, maternal education, socioeconomic status, gestational weight gain, and maternal diabetes mellitus status. NS (not significant): $p > 0.05$ ; \* $p \leq 0.05$ ; \*\* $p \leq 0.01$ ; \*\*\* $p \leq 0.001$ ; \*\*\*\* $p \leq 0.0001$ .

### Higher maternal BMI associated with MVM among term pregnancies

At term, increasing maternal BMI was associated with greater odds of an appropriately mature placenta based on apparent maturity of the placenta (aOR=0.94 [0.92–0.97]) and increased MVM (aβ=0.007 [0.004–0.01], Table 4), and placentae from mothers with obesity had the greatest percentage of thrombosed fetal vessels compared to all other BMI groups (Cramer’s V=0.03 [0.004–0.05], Supplementary Table 6). Additionally, presence of infarcts alone was more likely as maternal BMI increased (aOR=1.03 [1.02– 1.05], Supplementary Table 7). There were no differences in apparent placental maturity or vasculature-related pathologies by BMI among preterm pregnancies (Table 1).

### Both maternal underweight and increased BMI influenced placental anthropometry

In preterm pregnancies where mothers were underweight, placental weight was reduced compared to placentae from mothers with overweight and obesity (r=0.04 [-0.004 to 0.09]), and smallest diameter was reduced compared to placentae from pregnancies complicated by obesity (r=0.06 [0.01–0.11], Supplementary Table 8), but there were no differences in placental anthropometry compared to normal weight BMI. Among term pregnancies where mothers were underweight, placental weight (r=0.08 [0.06–0.10]) and smallest diameter (r=0.05 [0.03–0.07]) were reduced compared to all other BMI groups, with most prominent differences between the underweight and obese groups (Supplementary Table 6). At term only, birthweight to placental weight ratio was higher in mothers who were underweight compared to overweight, but not different than normal weight (r=-0.03 [-0.05 to -0.006], Supplementary Table 6). Similarly, when considering BMI as a continuous variable, placental weight and smallest diameter increased with increasing maternal BMI among preterm and term pregnancies, and birthweight to placental weight ratio decreased with increasing maternal BMI at term (Supplementary Tables 7,9).

### Fetal sex influenced placental pathology

Among preterm pregnancies, female placentae had a greater fetal inflammation summary score than male placentae (r=-0.05 [-0.10 to -0.004], Table 5), but there were no sex differences in neutrophil infiltration of the membranes or placental surface. Male term placentae had greater fetal and maternal placental inflammation, including neutrophil infiltration of the umbilical vein, umbilical cord substance, amnion membrane, chorion membrane, amnion of the placenta, chorion of the placenta, and fetal and maternal inflammation summary scores (Table 5). Further, when considering only pregnancies with maternal obesity, at term, male placentae had greater neutrophil infiltration of the umbilical vein and chorion membrane than female placentae (Supplementary Figure 3).

**Table 5.**
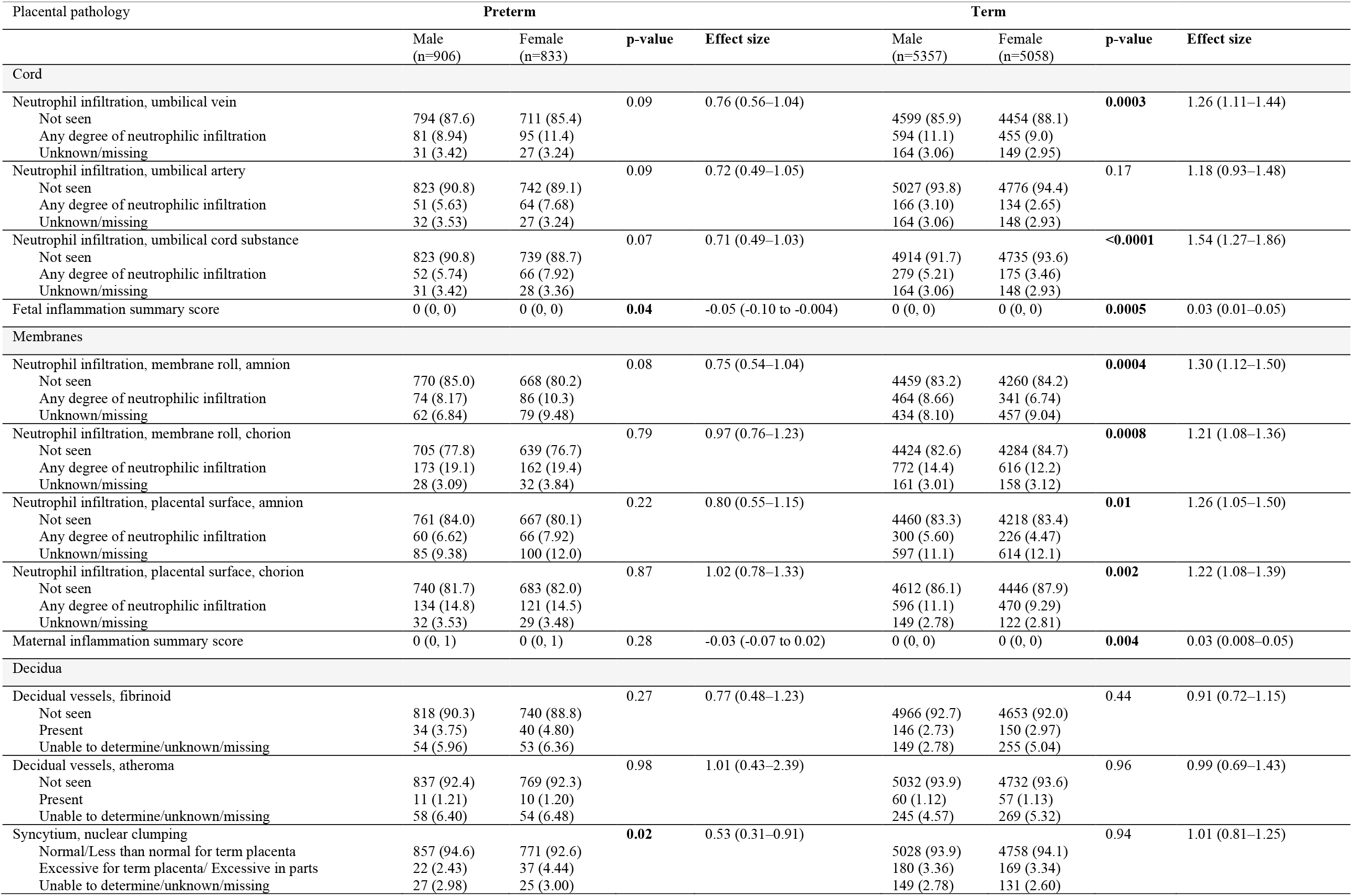
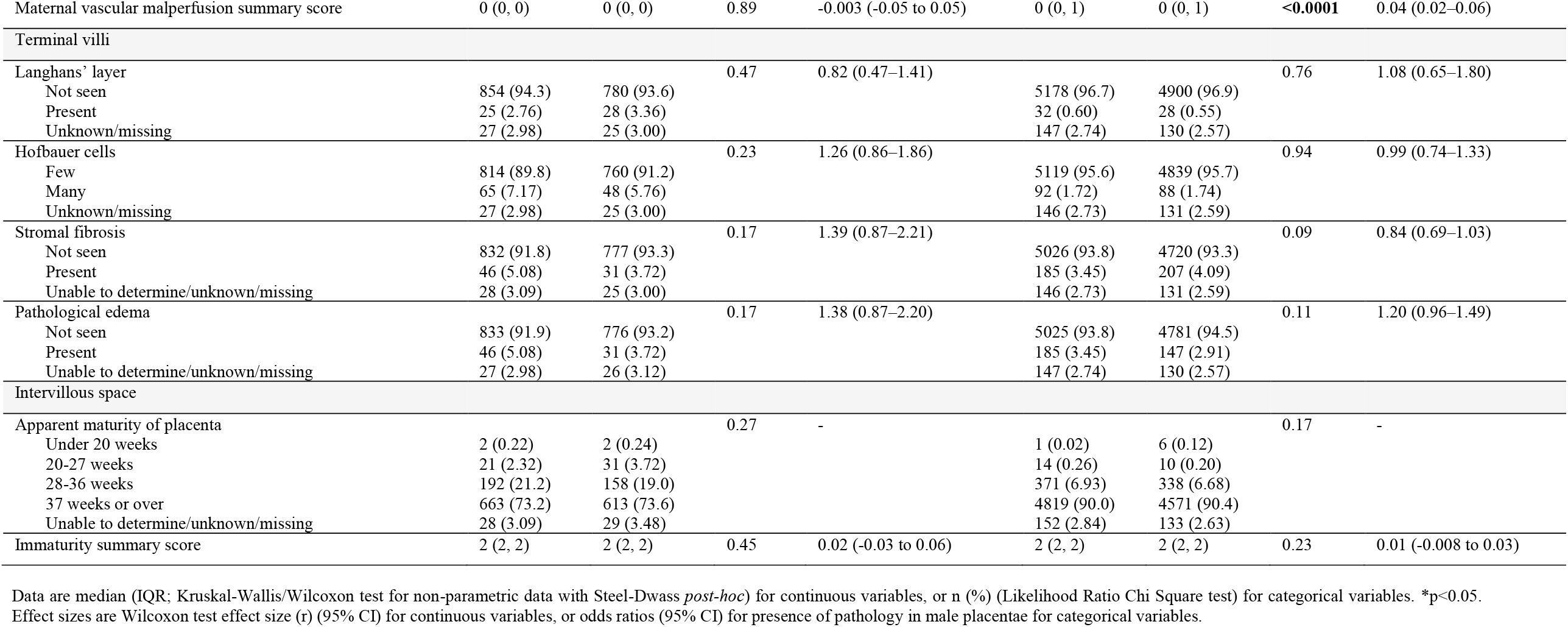
Associations between fetal sex and microscopic placental pathologies in preterm (N=1739) and term (N=10,415) pregnancies.

Female preterm placentae had increased syncytium-nuclear clumping (OR=0.53 [0.31– 0.91], Table 5), but there were no differences in presence of infarcts or MVM summary score between sexes preterm. Male term placentae had increased MVM (r=0.04 [0.02– 0.06], Table 5) and placental infarcts compared to female placentae (OR=1.22 [1.11– 1.33], Table 6). Infant birthweight to placental weight ratio was also increased in males compared to females at preterm (r=0.07 [0.03–0.12]) and term (r=0.07 [0.05–0.09], Table 6).

**Table 6.** Associations between fetal sex and macroscopic placental pathologies in preterm (N=1739) and term (N=10,415) pregnancies.

| Placental pathology | Preterm |  |  |  | Term |  |  |  |
| --- | --- | --- | --- | --- | --- | --- | --- | --- |
|  | Male<br>(n=906) | Female<br>(n=833) | p-value | Effect size | Male<br>(n=5357) | Female<br>(n=5058) | p-value | Effect size |
| Placenta<sup>1</sup> |  |  |  |  |  |  |  |  |
| Placenta shape |  |  | 0.64 | - |  |  | 0.17 | - |
| Non-partite | 804 (88.7) | 727 (87.3) |  |  | 4839 (90.3) | 4553 (90.0) |  |  |
| Bipartite | 4 (0.44) | 1 (0.12) |  |  | 65 (1.21) | 68 (1.34) |  |  |
| Tripartite | 0 | 0 |  |  | 0 | 2 (0.04) |  |  |
| Succenturiate | 12 (1.32) | 11 (1.32) |  |  | 61 (1.14) | 53 (1.05) |  |  |
| Membranous placenta | 0 | 0 |  |  | 0 | 2 (0.04) |  |  |
| Crescent shaped | 0 | 0 |  |  | 2 (0.04) | 0 |  |  |
| Irregular shape | 4 (0.44) | 3 (0.36) |  |  | 23 (0.43) | 23 (0.45) |  |  |
| Unknown/missing | 82 (9.05) | 91 (10.9) |  |  | 518 (9.67) | 357 (7.06) |  |  |
| Weight | 382 (320, 450) | 375 (320, 435) | 0.17 | 0.03 (-0.02 to 0.08) | 425 (370, 485) | 420 (370, 480) | 0.06 | 0.02 (-0.002 to 0.04) |
| Largest diameter | 18 (17, 19) | 18 (17, 19) | 0.61 | 0.01 (-0.04 to 0.06) | 19 (18, 20) | 19 (18, 20) | **0.03** | 0.02 (0.002–0.04) |
| Smallest diameter | 16 (14, 17) | 15 (14, 17) | 0.41 | 0.02 (-0.03 to 0.07) | 16 (15, 18) | 16 (15, 17) | 0.19 | 0.01 (-0.006 to 0.03) |
| Thickness | 20 (18, 24) | 20 (18, 23) | 0.38 | 0.02 (-0.02 to 0.07) | 20 (20, 25) | 20 (20, 25) | 0.53 | 0.006 (-0.02 to 0.03) |
| Birthweight : placenta weight | 7.34 (6.52, 8.32) | 7.16 (6.36, 7.93) | **0.003** | 0.07 (0.03–0.12) | 7.56 (6.78, 8.38) | 7.36 (6.65, 8.17) | **<0.0001** | 0.07 (0.05–0.09) |
| Birthweight : placental largest diameter | 157 (140, 175) | 150 (135, 166) | **<0.0001** | 0.13 (0.09–0.018) | 169 (155, 184) | 165 (151, 179) | **<0.0001** | 0.09 (0.07–0.11) |
| Birthweight : placental smallest diameter | 183 (164, 202) | 173 (156, 193) | **<0.0001** | 0.12 (0.08–0.17) | 197 (179, 214) | 191 (175, 208) | **<0.0001** | 0.09 (0.07–0.11) |
| Birthweight : placental thickness | 140 (118, 162) | 133 (116, 155) | **0.003** | 0.07 (0.03–0.12) | 147 (126, 170) | 143 (125, 164) | **<0.0001** | 0.06 (0.04–0.08) |
| Cord |  |  |  |  |  |  |  |  |
| Cord number of vessels |  |  | 0.85 | 0.90 (0.29–2.79) |  |  | 0.10 | 0.66 (0.40–1.09) |
| Two | 821 (90.6) | 736 (88.4) |  |  | 4965 (92.7) | 4659 (92.1) |  |  |
| Three | 6 (0.66) | 6 (0.72) |  |  | 26 (0.49) | 37 (0.73) |  |  |
| Other/unknown/missing | 79 (8.72) | 91 (10.9) |  |  | 366 (6.83) | 362 (7.16) |  |  |
| Cord edema |  |  | 0.97 | 1.01 (0.62–1.64) |  |  | **0.02** | 1.46 (1.07–1.99) |
| Not seen | 781 (86.2) | 700 (84.0) |  |  | 4817 (89.9) | 4562 (90.2) |  |  |
| Present | 36 (3.97) | 31 (3.72) |  |  | 103 (1.92) | 67 (1.32) |  |  |
| Unknown/missing | 89 (9.82) | 101 (12.1) |  |  | 437 (8.16) | 429 (8.48) |  |  |
| Membranes and fetal surface |  |  |  |  |  |  |  |  |
| Opacity of membranes |  |  | 0.97 | 1.01 (0.69–1.46) |  |  | 0.45 | 1.06 (0.91–1.25) |
| Not opaque | 763 (84.2) | 684 (82.1) |  |  | 4648 (86.8) | 4384 (86.7) |  |  |
| Any degree of opacity | 64 (7.06) | 57 (6.84) |  |  | 347 (6.48) | 308 (6.09) |  |  |
| Unknown/missing | 79 (8.72) | 92 (11.0) |  |  | 362 (6.76) | 366 (7.24) |  |  |
| Thrombosed fetal vessels |  |  | 0.93 | 1.05 (0.35–3.13) |  |  | 0.73 | 0.88 (0.42–1.82) |
| Not seen | 821 (90.6) | 738 (88.6) |  |  | 4983 (93.0) | 4687 (92.7) |  |  |
| Present | 7 (0.77) | 6 (0.72) |  |  | 14 (0.26) | 15 (0.30) |  |  |
| Unknown/missing | 78 (8.61) | 89 (10.7) |  |  | 360 (6.72) | 356 (7.04) |  |  |
| Cut surface |  |  |  |  |  |  |  |  |
| Infarct size |  |  | 0.89 | 0.98 (0.77–1.26) |  |  | **<0.0001** | 1.22 (1.11–1.33) |
| Not applicable | 660 (72.8) | 593 (71.2) |  |  | 3546 (66.2) | 3513 (69.5) |  |  |
| Any infarcts seen | 164 (18.1) | 150 (18.0) |  |  | 1433 (26.8) | 1167 (23.1) |  |  |
| Unknown/missing | 82 (9.05) | 90 (10.8) |  |  | 378 (7.06) | 378 (7.47) |  |  |
| Cut surface calcification |  |  | 0.43 | 1.08 (0.89–1.32) |  |  | 0.14 | 0.94 (0.87–1.02) |
| None | 444 (49.0) | 414 (49.7) |  |  | 2051 (66.2) | 1860 (36.8) |  |  |
| Any calcification seen | 380 (41.9) | 327 (39.3) |  |  | 2926 (54.6) | 2819 (55.7) |  |  |
| Unknown/missing | 82 (9.05) | 92 (11.0) |  |  | 380 (7.09) | 379 (7.49) |  |  |

Data are median (IQR; Kruskal-Wallis/Wilcoxon test for non-parametric data with Steel-Dwass *post-hoc*) for continuous variables, or n (%) (Likelihood Ratio Chi Square test) for categorical variables. \*p<0.05. Effect sizes are Wilcoxon test effect size (r) (95% CI) for continuous variables, or odds ratios (95% CI) for presence of pathology in male placentae for categorical variables. <sup>1</sup>The top and bottom 0.5% of raw infant and placental anthropometry data (birthweight [g], placental weight [g], placental diameter [cm], and placental thickness [mm]) were excluded to remove biologically implausible data.

## Discussion

We evaluated the associations between maternal prepregnancy BMI and the prevalence of placental pathologies in preterm and term pregnancies, to better understand the placental mechanisms that may explain poor pregnancy and offspring outcomes in pregnancies complicated by suboptimal maternal BMI. Using data from 12,154 pregnancies from the Collaborative Perinatal Project, we found placental inflammation was increased in pregnancies complicated by maternal obesity, and notably, the inflammatory response was different based on gestational age. Maternal obesity also associated with increased MVM of the placenta, and increased maternal BMI associated with greater odds of having an appropriate mature placenta at term. Placental efficiency was highest in pregnancies where mothers were underweight and in male placentae, and pathologies also differed by fetal sex. Suboptimal maternal BMI thus alters normal gestational tissue development, with likely effects on function.

The inflammatory conditions established by maternal obesity may favour placental inflammation. In support of this, we found that higher maternal BMI associated with increased fetal (e.g. neutrophilic infiltration of the umbilical vein, artery, and cord substance) inflammation at preterm, and maternal (e.g. neutrophilic infiltration of the amnion and chorion membranes, amnion and chorion of the placenta, and opacity of membranes) gestational tissue inflammation at term. While few studies have characterised fetal- and maternal-specific placental inflammation in the context of both maternal BMI and gestational age, at term, maternal inflammation has been previously characterized in pregnancies with obesity, evidenced by increased pro-inflammatory cytokines in the placenta^30^, and increased maternal, but not fetal, inflammatory lesions^17,56^. Other studies have found no differences in placental inflammation by maternal BMI in term pregnancies, however, these cohorts had a small number of cases^57,58^. Less is known about the relationships between maternal obesity and specific placental inflammation at preterm, however, maternal obesity has been associated with greater risk of chorioamnionitis leading to preterm birth^59^, and a combination of maternal (defined as inflammation in the chorion, amnion, and decidua) and fetal (inflammation of the umbilical cord and chorionic plate fetal vessels) gestational tissue inflammation has been linked to higher risk of extreme preterm birth than maternal gestational tissue inflammation alone^60^. In the context of ascending infection, fetal placental inflammation of the umbilical cord has been associated with greater neonatal morbidity and mortality, particularly among preterm pregnancies, than maternal placental inflammation^61-63^. Our study, unlike many others, considers both suboptimal maternal BMI and gestational age, and suggests that increased maternal BMI associates with fetal placental inflammation at preterm, which has been linked to adverse pregnancy and offspring outcomes^64,65^, and maternal focused placental inflammation at term. Fetal inflammation of the placenta at preterm could thus be an important risk factor for adverse offspring outcomes, and maternal placental inflammation, though less frequently associated with clinical correlates^64^, nonetheless suggests that suboptimal maternal BMI alters gestational tissue histomorphology.

Interestingly, preterm female placentae had increased fetal inflammation, while term male placentae had increased fetal and maternal inflammation. Few studies have observed gestational age-specific sex differences in placental inflammation, and studies that have shown increased female gestational tissue inflammation often only consider term placentae. For example, female placentae have been shown to have increased chronic villitis in pregnancies complicated by maternal obesity, however those observations were restricted to pregnancies at or near term^15^, as well as increased expression of genes related to immune regulation in normal, term pregnancies^66^. Sex-specific differences have also been observed in pregnancies exacerbated by other inflammatory conditions, where female placentae have increased mRNA expression levels of TNF-α, IL-1β, IL-6, IL-8, and IL-5 in pregnancies where mothers had asthma, compared to levels in male placentae^67^. Further, peripheral blood mononuclear cells treated with lipopolysaccharide from pregnant women carrying female fetuses had greater stimulated production of IL-6, TNF-α, and IL-1β compared to women carrying male feutses, suggesting a greater response to immune challenge^68^. These findings may suggest that male and female fetuses implement different survival strategies in response to the same adverse environmental stimuli, where females may mount stronger responses, while males make minimal adaptations and continue to prioritize growth^69,70^. This may explain our finding of increased female fetal gestational tissue inflammation. Further, smaller body size^71,72^ may allow female fetuses to gestate longer in the context of intrauterine inflammation, while continuing to mount an immune response to adverse environmental conditions, while larger male fetuses^71,72^ may trigger uterine mechanical stretch, and in conjunction with inflammatory signals^73,74^, initiate earlier parturition.

In contrast, male placentae have been observed to have higher rates of chronic inflammatory lesions in extreme preterm pregnancies^75^, chronic deciduitis among extreme preterm pregnancies with pre-eclampsia and intrauterine growth restriction^76^, and enrichment in inflammatory pathways^77^. Our findings suggest that male placentae are more susceptible to inflammation in term pregnancies, which could increase risk for impaired growth or neurodevelopment and long term health outcomes^78,79^, while increased fetal inflammation among preterm female placentae warrants further investigation. Given the associations of maternal obesity and male sex with increased placental inflammation, term males born to mothers with obesity may be most susceptible to placental inflammation. To further investigate this, we conducted an exploratory analysis to assess whether fetal sex associated with placental inflammation among pregnancies with obesity alone. Increased odds of inflammation in male placentae at term only, though not to the same extent as BMI inclusive differences, suggests that both maternal obesity and male sex may be independent risk factors for placental inflammation at term. Further studies are needed to corroborate this finding.

We found MVM also associated with increased maternal BMI among term pregnancies, consistent with existing data^80^. Previous studies have documented increased maternal vascular lesions^58^ and decidual vasculopathy, but no other lesions associated with MVM, in placentae from term pregnancies complicated by obesity compared to normal weight, although this was attributed to maternal hypertensive disease^15^. MVM lesions impair intervillous blood flow, altering oxygen and nutrient delivery to the fetus^47^, and are associated with adverse offspring outcomes, including preterm birth, intrauterine growth restriction, and small for gestational age infants^47^. Our findings suggest that the adverse environment established by maternal obesity may impair the development and function of placental vasculature, and thus fetal development. Despite known associations of MVM and preterm birth^47,81^, increased BMI associated with MVM only among term pregnancies in our cohort^15,47,58^, suggesting the importance of vascular pathologies across gestation in pregnancies complicated by maternal obesity. Further, male placentae had increased MVM lesions compared to females at term in our study, consistent with other findings of increased decidual vasculopathy in males^15^. Thus, placental vasculature pathology may not be so severe as to result in early pregnancy or other adverse perinatal events, but may still have long term implications for the infant.

In contrast to previous studies, we found increased maternal BMI associated with greater odds of having an appropriately mature placenta at term. Placentae from women with obesity have been shown to have immaturity of the villous tree compared to women of normal weight^57^, suggesting structural and functional maladaptation of the vasculature or decreased efficiency in maternal-fetal exchange^57^. We did, however observe other placental vascular pathologies, evidenced by increased MVM with higher maternal BMI. Placental maturity was classified by CPP pathologists as placental appearance of <20, 20–27, 28–36, or ≥37 weeks’ gestation, based on presence of fibrin under the chorionic plate, presence of cysts on the cut surfaces, lack of Langhans layer, relative uniformity of villous size, crowded fetal capillaries within villi, and increased frequency of syncytial knots. In addition to the lower number of cases of obesity in our cohort relative to the current rates, changes to criteria used to diagnose distal villous immaturity, which was only introduced after the time of the CPP^82^, may in part account for these differences in apparent placental maturity. Overall, a consistent, universal classification of placental maturation disorders would be beneficial in reducing variation in assessment of placental maturity across studies^83^.

Our findings of decreased placental size and increased infant birthweight to placental weight ratio with lower maternal BMI are consistent with reduced nutrient availability in underweight pregnancies^5,20,84^, and suggest that the placenta may adapt to increase nutrient delivery to the fetus, whereas placentae from higher BMI pregnancies may adapt by regulating nutrient transfer to the fetus in the face of sufficient or overabundance of nutrients^85,86^. Conversely, among term pregnancies, decreased birthweight to placental smallest diameter ratio with lower maternal BMI could suggest lower placental efficiency in pregnancies with underweight. However, placental diameter is reflective of the lateral growth of the placenta and area of the uterine lining that it encompasses, and may be influenced by inter-individual variations such as differences in placental shape, whereas placental weight captures multiple dimensions of placental growth^87^. Further, males had higher birthweight to placental size ratios in both preterm and term pregnancies, consistent with previous studies^88^, suggesting greater placental efficiency than females. Male fetuses tend to grow more rapidly than females and invest greater resources in growth than placental development and reserve capacity^66,69-71^, and as a result, may be more susceptible to placental insults as well as adverse later health outcomes^71^. Sex-based differences in fetoplacental growth and development may be attributed to a number of contributing biological factors, including X chromosome inactivation^69,89^, sex-specific response to glucocorticoids^69,90^, and levels of sex hormones^69^. Taken together, our results may suggest that males born to mothers who are underweight have the greatest placental efficiency, particularly at term.

Strengths of our study include the large population-based cohort, where previous studies have been limited to animal models of maternal malnutrition or inflammation to assess placental histopathology, or have lacked population size and comprehensive data. Importantly, the large sample size, collection of socioeconomic and demographic factors, deep phenotyping, and prospective nature of the dataset enables a thorough investigation of the relationships between maternal BMI and placental pathologies in a diverse population. To leverage the extensive data collected, future studies could further explore the role of demographic factors on placental pathology, such as socioeconomic status, in addition to suboptimal maternal BMI, given the independent risks associated with both low socioeconomic status^91^ and with maternal underweight and obesity^15-17,19^ on outcomes such as preterm birth and placental pathology, respectively. Due to the cohort’s historical nature, a limitation of our study is that the prevalence of obesity was lower than current rates; only 2.63% of our cohort were classified as having obesity prepregnancy, compared to 29% of American women currently^92^. Rates of smoking were also increased during the time of the study compared to rates today (42-45% during the time of CPP data collection^93^ compared to 16% in 2019^94^), and no data were collected on maternal alcohol consumption^95^. Additionally, no data were provided for GDM specifically, which is associated with adverse pregnancy outcomes and is comorbid with obesity^96,97^, however, rates of GDM were much lower during the time of the study than current rates (0.3% in 1979^98^ compared to 7.6% from 2007-2014^99^), so any confounding is likely to be minimal. Placental pathology was not re-assessed, however, definitions of pathologies were largely consistent between the CPP and current Amsterdam Placental Workshop Group Consensus Statement criteria^18^, particularly for our variables of interest. Given the population-based design, our findings may still be applicable to a broad and diverse population today, due to the large sample size and thorough collection of demographic and placental pathology data.

Given our findings of suboptimal maternal BMI, particularly high BMI, and risk of placental pathology, strategies to optimize maternal weight during pregnancy that align with improving patient-centred health outcomes and addressing root causes of obesity^100^ should be investigated for their effects on placental development and function, including other pregnancy outcomes. To date, there is limited evidence that dietary and lifestyle interventions are effective in improving pregnancy outcomes for women who have high BMI, but may still have implications for weight management. For example, a diet and physical activity intervention improved maternal lipid metabolic profiles across pregnancy in women with obesity, but did not improve the primary study outcomes of interest, prevalence of maternal diabetes mellitus and large for gestational age infants at birth^101^. However, in addition to improved diet and physical activity, the trial did show beneficial changes in maternal gestational weight gain^101^, and similar improvements in gestational weight gain have been seen in other diet and physical activity studies among women who were overweight prepregnancy^102^. Nonetheless, it is possible that modest changes in maternal diet and activity may result in subtle, but important, improvements in placental development and function, measures not often considered in maternal lifestyle intervention studies. For example, maternal exercise in a high fat diet mouse model of maternal obesity has shown improvements in maternal weight gain, serum glucose and lipid levels, and insulin sensitivity, and protective effects on placental vascular factors, in addition to prevented fetal overgrowth^103^. This study contributes to better understanding the role of the placenta in pregnancies complicated by suboptimal maternal BMI, to ultimately help inform clinical and policy guidelines.

Our data demonstrate that compared to normal weight, maternal underweight and obesity prepregnancy, even in the absence of other significant pregnancy complications, are not inert conditions for the developing placenta^16,37^, which may have consequences not only for immediate pregnancy and fetal outcomes but postnatal growth and health trajectories^38,39,44,104^. Characterising placental (mal)adaptations to common maternal conditions using clinically-relevant indicators can help to understand the mechanisms through which these conditions affect the developing offspring, and aid clinical decision making to better support high-risk pregnancies and inform interventions to optimise pregnancy, placental, and infant health.

## Data Availability

The data that support the findings of this study are available in the United States National Archives at https://www.archives.gov/research/electronic-records/nih.html, National Archives Identifier: 606622 (Record Group 443: Records of the National Institutes of Health [NIH]). Derived data supporting the findings of this study are available from the corresponding author on reasonable request.

## Funding and Competing Interests

The authors have no competing interests to declare. This research is funded by the Faculty of Science, Carleton University. HS was supported by a Mitacs Research Training Award. KLC is supported by grants from the Canadian Institutes of Health Research, Natural Sciences and Engineering Research Council of Canada, the Molly Towell Perinatal Research Foundation (New Investigator), and Carleton University Office of Research.

### Acknowledgments

The authors thank the U.S. National Archives for the publicly available Collaborative Perinatal Project data, and Marina White for her assistance with obtaining the dataset.

## Author Contributions

Conceptualization, methodology: HS, KLC, DG, LNA; formal analysis: HS; writing—original draft preparation: HS, KLC; writing—review and editing: HS, KLC, DG, LNA; visualization: HS, KLC.

## Supplementary tables

**Supplementary Table 1.** Binary categories for placental pathology variables for multivariable analysis.

| Categorical variable | Collapsed binary categories |
| --- | --- |
| Cord number of vessels | Three vessels vs two vessels |
| Cord edema<br>(derived from “cord description” variable) | Presence of cord edema vs absence of cord edema |
| Opacity of membranes | Not opaque vs any degree of opacity (partially opaque, opaque) |
| Thrombosed fetal vessel | Not seen vs present |
| Placental infarcts<br>(derived from “infarcts size” variable) | No infarcts seen (not applicable) vs any infarcts seen (all infarcts less than three centimetres, at least one infarct measures three or more centimetres) |
| Cut surface calcification | Not seen (none) vs any calcification seen (maternal surface only, calcification throughout) |
| Neutrophilic infiltration | Not seen vs any degree of neutrophilic infiltration seen (slight, moderate, or marked) |
| Decidual vessels fibrinoid | Not seen vs present |
| Decidual vessels atheroma | Not seen vs present |
| Langhan’s layer | Not seen vs present |
| Syncytium nuclear clumping | Normal/less than normal for term placenta vs excessive for term placenta/excessive in parts |
| Hofbauer cells | Few vs many |
| Stromal fibrosis | Not seen vs present (present- not qualified further, present- rare, present- many) |
| Pathological edema | Not seen vs present (present- not qualified further, present- rare, present- many) |
| Apparent maturity of the placenta | Preterm pregnancies: appropriate mature ($\leq 36$ weeks) vs hyper-mature ($\geq 37$ weeks)<br>Term pregnancies: appropriate mature ($\geq 37$ weeks) vs immature ($\leq 36$ weeks) |

**Supplementary Table 2.**
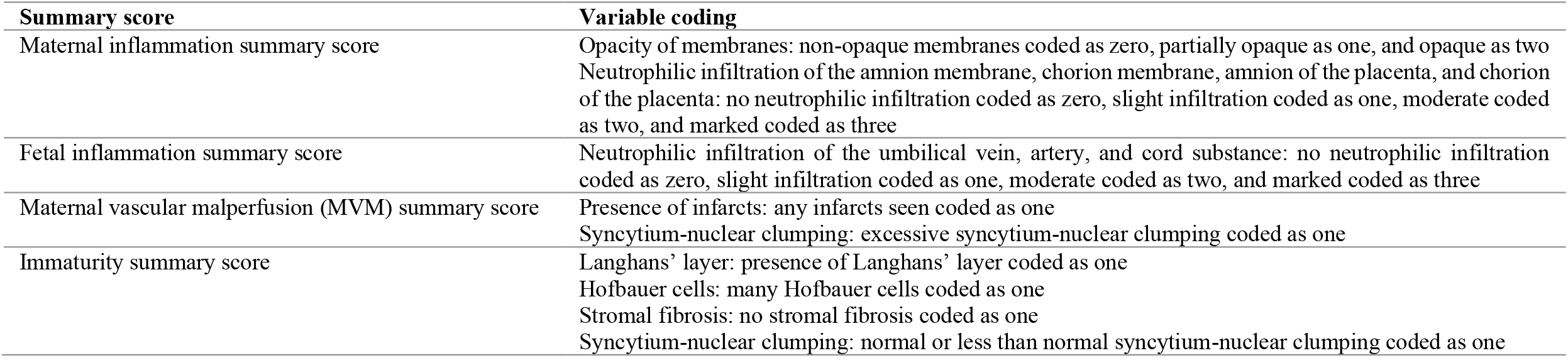
Derived summary scores variable coding.

| Summary score | Variable coding |
| --- | --- |
| Maternal inflammation summary score | Opacity of membranes: non-opaque membranes coded as zero, partially opaque as one, and opaque as two<br>Neutrophilic infiltration of the amnion membrane, chorion membrane, amnion of the placenta, and chorion of the placenta: no neutrophilic infiltration coded as zero, slight infiltration coded as one, moderate coded as two, and marked coded as three |
| Fetal inflammation summary score | Neutrophilic infiltration of the umbilical vein, artery, and cord substance: no neutrophilic infiltration coded as zero, slight infiltration coded as one, moderate coded as two, and marked coded as three |
| Maternal vascular malperfusion (MVM) summary score | Presence of infarcts: any infarcts seen coded as one<br>Syncytium-nuclear clumping: excessive syncytium-nuclear clumping coded as one |
| Immaturity summary score | Langhans' layer: presence of Langhans' layer coded as one<br>Hofbauer cells: many Hofbauer cells coded as one<br>Stromal fibrosis: no stromal fibrosis coded as one<br>Syncytium-nuclear clumping: normal or less than normal syncytium-nuclear clumping coded as one |

**Supplementary Table 3.**
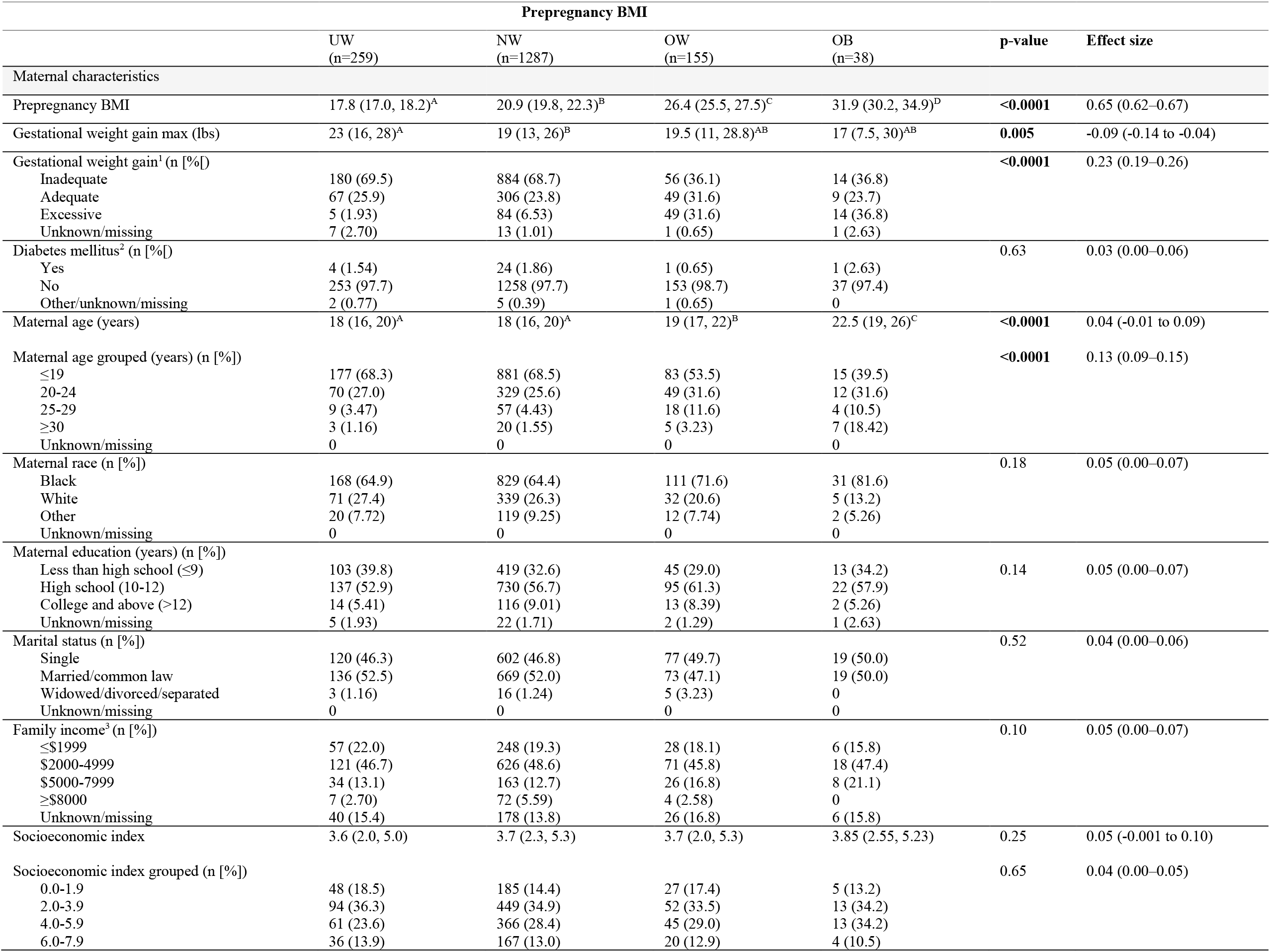
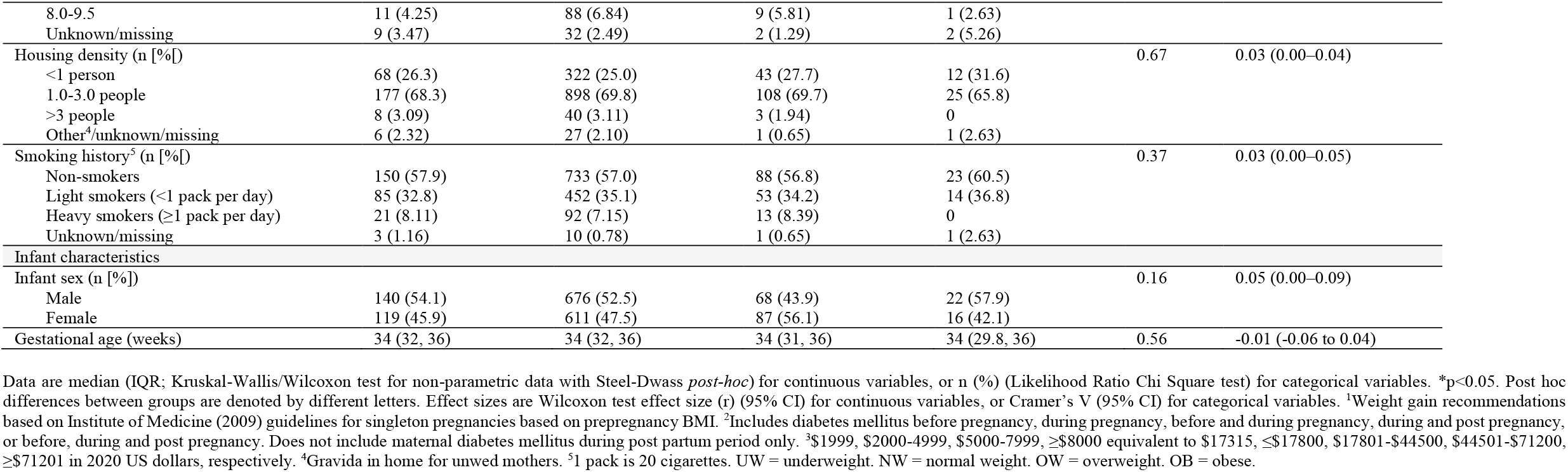
Maternal characteristics by prepregnancy BMI in preterm pregnancies, N=1739.

|  | Prepregnancy BMI |  |  |  |  |  |
| --- | --- | --- | --- | --- | --- | --- |
|  | UW<br>(n=259) | NW<br>(n=1287) | OW<br>(n=155) | OB<br>(n=38) | p-value | Effect size |
| Maternal characteristics |  |  |  |  |  |  |
| Pregpregnancy BMI | 17.8 (17.0, 18.2)<sup>A</sup> | 20.9 (19.8, 22.3)<sup>B</sup> | 26.4 (25.5, 27.5)<sup>C</sup> | 31.9 (30.2, 34.9)<sup>D</sup> | <0.0001 | 0.65 (0.62–0.67) |
| Gestational weight gain max (lbs) | 23 (16, 28)<sup>A</sup> | 19 (13, 26)<sup>B</sup> | 19.5 (11, 28.8)<sup>AB</sup> | 17 (7.5, 30)<sup>AB</sup> | 0.005 | -0.09 (-0.14 to -0.04) |
| Gestational weight gain<sup>1</sup> (n [%]) |  |  |  |  | <0.0001 | 0.23 (0.19–0.26) |
| Inadequate | 180 (69.5) | 884 (68.7) | 56 (36.1) | 14 (36.8) |  |  |
| Adequate | 67 (25.9) | 306 (23.8) | 49 (31.6) | 9 (23.7) |  |  |
| Excessive | 5 (1.93) | 84 (6.53) | 49 (31.6) | 14 (36.8) |  |  |
| Unknown/missing | 7 (2.70) | 13 (1.01) | 1 (0.65) | 1 (2.63) |  |  |
| Diabetes mellitus<sup>2</sup> (n [%]) |  |  |  |  | 0.63 | 0.03 (0.00–0.06) |
| Yes | 4 (1.54) | 24 (1.86) | 1 (0.65) | 1 (2.63) |  |  |
| No | 253 (97.7) | 1258 (97.7) | 153 (98.7) | 37 (97.4) |  |  |
| Other/unknown/missing | 2 (0.77) | 5 (0.39) | 1 (0.65) | 0 |  |  |
| Maternal age (years) | 18 (16, 20)<sup>A</sup> | 18 (16, 20)<sup>A</sup> | 19 (17, 22)<sup>B</sup> | 22.5 (19, 26)<sup>C</sup> | <0.0001 | 0.04 (-0.01 to 0.09) |
| Maternal age grouped (years) (n [%]) |  |  |  |  | <0.0001 | 0.13 (0.09–0.15) |
| ≤19 | 177 (68.3) | 881 (68.5) | 83 (53.5) | 15 (39.5) |  |  |
| 20-24 | 70 (27.0) | 329 (25.6) | 49 (31.6) | 12 (31.6) |  |  |
| 25-29 | 9 (3.47) | 57 (4.43) | 18 (11.6) | 4 (10.5) |  |  |
| ≥30 | 3 (1.16) | 20 (1.55) | 5 (3.23) | 7 (18.42) |  |  |
| Unknown/missing | 0 | 0 | 0 | 0 |  |  |
| Maternal race (n [%]) |  |  |  |  | 0.18 | 0.05 (0.00–0.07) |
| Black | 168 (64.9) | 829 (64.4) | 111 (71.6) | 31 (81.6) |  |  |
| White | 71 (27.4) | 339 (26.3) | 32 (20.6) | 5 (13.2) |  |  |
| Other | 20 (7.72) | 119 (9.25) | 12 (7.74) | 2 (5.26) |  |  |
| Unknown/missing | 0 | 0 | 0 | 0 |  |  |
| Maternal education (years) (n [%]) |  |  |  |  | 0.14 | 0.05 (0.00–0.07) |
| Less than high school (≤9) | 103 (39.8) | 419 (32.6) | 45 (29.0) | 13 (34.2) |  |  |
| High school (10-12) | 137 (52.9) | 730 (56.7) | 95 (61.3) | 22 (57.9) |  |  |
| College and above (>12) | 14 (5.41) | 116 (9.01) | 13 (8.39) | 2 (5.26) |  |  |
| Unknown/missing | 5 (1.93) | 22 (1.71) | 2 (1.29) | 1 (2.63) |  |  |
| Marital status (n [%]) |  |  |  |  | 0.52 | 0.04 (0.00–0.06) |
| Single | 120 (46.3) | 602 (46.8) | 77 (49.7) | 19 (50.0) |  |  |
| Married/common law | 136 (52.5) | 669 (52.0) | 73 (47.1) | 19 (50.0) |  |  |
| Widowed/divorced/separated | 3 (1.16) | 16 (1.24) | 5 (3.23) | 0 |  |  |
| Unknown/missing | 0 | 0 | 0 | 0 |  |  |
| Family income<sup>3</sup> (n [%]) |  |  |  |  | 0.10 | 0.05 (0.00–0.07) |
| ≤\$1999 | 57 (22.0) | 248 (19.3) | 28 (18.1) | 6 (15.8) | | |
| \$2000-4999 | 121 (46.7) | 626 (48.6) | 71 (45.8) | 18 (47.4) | | |
| \$5000-7999 | 34 (13.1) | 163 (12.7) | 26 (16.8) | 8 (21.1) | | |
| ≥\$8000 | 7 (2.70) | 72 (5.59) | 4 (2.58) | 0 | | |
| Unknown/missing | 40 (15.4) | 178 (13.8) | 26 (16.8) | 6 (15.8) |  |  |
| Socioeconomic index | 3.6 (2.0, 5.0) | 3.7 (2.3, 5.3) | 3.7 (2.0, 5.3) | 3.85 (2.55, 5.23) | 0.25 | 0.05 (-0.001 to 0.10) |
| Socioeconomic index grouped (n [%]) |  |  |  |  | 0.65 | 0.04 (0.00–0.05) |
| 0.0-1.9 | 48 (18.5) | 185 (14.4) | 27 (17.4) | 5 (13.2) |  |  |
| 2.0-3.9 | 94 (36.3) | 449 (34.9) | 52 (33.5) | 13 (34.2) |  |  |
| 4.0-5.9 | 61 (23.6) | 366 (28.4) | 45 (29.0) | 13 (34.2) |  |  |
| 6.0-7.9 | 36 (13.9) | 167 (13.0) | 20 (12.9) | 4 (10.5) |  |  |
| 8.0-9.5 | 11 (4.25) | 88 (6.84) | 9 (5.81) | 1 (2.63) |  |  |
| Unknown/missing | 9 (3.47) | 32 (2.49) | 2 (1.29) | 2 (5.26) |  |  |
| Housing density (n [%]) |  |  |  |  | 0.67 | 0.03 (0.00–0.04) |
| <1 person | 68 (26.3) | 322 (25.0) | 43 (27.7) | 12 (31.6) |  |  |
| 1.0-3.0 people | 177 (68.3) | 898 (69.8) | 108 (69.7) | 25 (65.8) |  |  |
| >3 people | 8 (3.09) | 40 (3.11) | 3 (1.94) | 0 |  |  |
| Other<sup>4</sup>/unknown/missing | 6 (2.32) | 27 (2.10) | 1 (0.65) | 1 (2.63) |  |  |
| Smoking history<sup>5</sup> (n [%]) |  |  |  |  | 0.37 | 0.03 (0.00–0.05) |
| Non-smokers | 150 (57.9) | 733 (57.0) | 88 (56.8) | 23 (60.5) |  |  |
| Light smokers (<1 pack per day) | 85 (32.8) | 452 (35.1) | 53 (34.2) | 14 (36.8) |  |  |
| Heavy smokers (≥1 pack per day) | 21 (8.11) | 92 (7.15) | 13 (8.39) | 0 |  |  |
| Unknown/missing | 3 (1.16) | 10 (0.78) | 1 (0.65) | 1 (2.63) |  |  |
| Infant characteristics |  |  |  |  |  |  |
| Infant sex (n [%]) |  |  |  |  | 0.16 | 0.05 (0.00–0.09) |
| Male | 140 (54.1) | 676 (52.5) | 68 (43.9) | 22 (57.9) |  |  |
| Female | 119 (45.9) | 611 (47.5) | 87 (56.1) | 16 (42.1) |  |  |
| Gestational age (weeks) | 34 (32, 36) | 34 (32, 36) | 34 (31, 36) | 34 (29.8, 36) | 0.56 | -0.01 (-0.06 to 0.04) |

Data are median (IQR; Kruskal-Wallis/Wilcoxon test for non-parametric data with Steel-Dwass *post-hoc* for continuous variables, or n (%) (Likelihood Ratio Chi Square test) for categorical variables. \*p<0.05. Post hoc differences between groups are denoted by different letters. Effect sizes are Wilcoxon test effect size (r) (95% CI) for continuous variables, or Cramer's V (95% CI) for categorical variables. <sup>1</sup>Weight gain recommendations based on Institute of Medicine (2009) guidelines for singleton pregnancies based on prepregnancy BMI. <sup>2</sup>Includes diabetes mellitus before pregnancy, during pregnancy, before and during pregnancy, during and post pregnancy, or before, during and post pregnancy. Does not include maternal diabetes mellitus during post partum period only. <sup>3</sup>\$1999, \$2000-4999, \$5000-7999, ≥\$8000 equivalent to \$17315, ≤\$17800, \$17801-\$44500, \$44501-\$71200, ≥\$71201 in 2020 US dollars, respectively. <sup>4</sup>Gravida in home for unwed mothers. <sup>5</sup>1 pack is 20 cigarettes. UW = underweight. NW = normal weight. OW = overweight. OB = obese.

**Supplementary Table 4.**
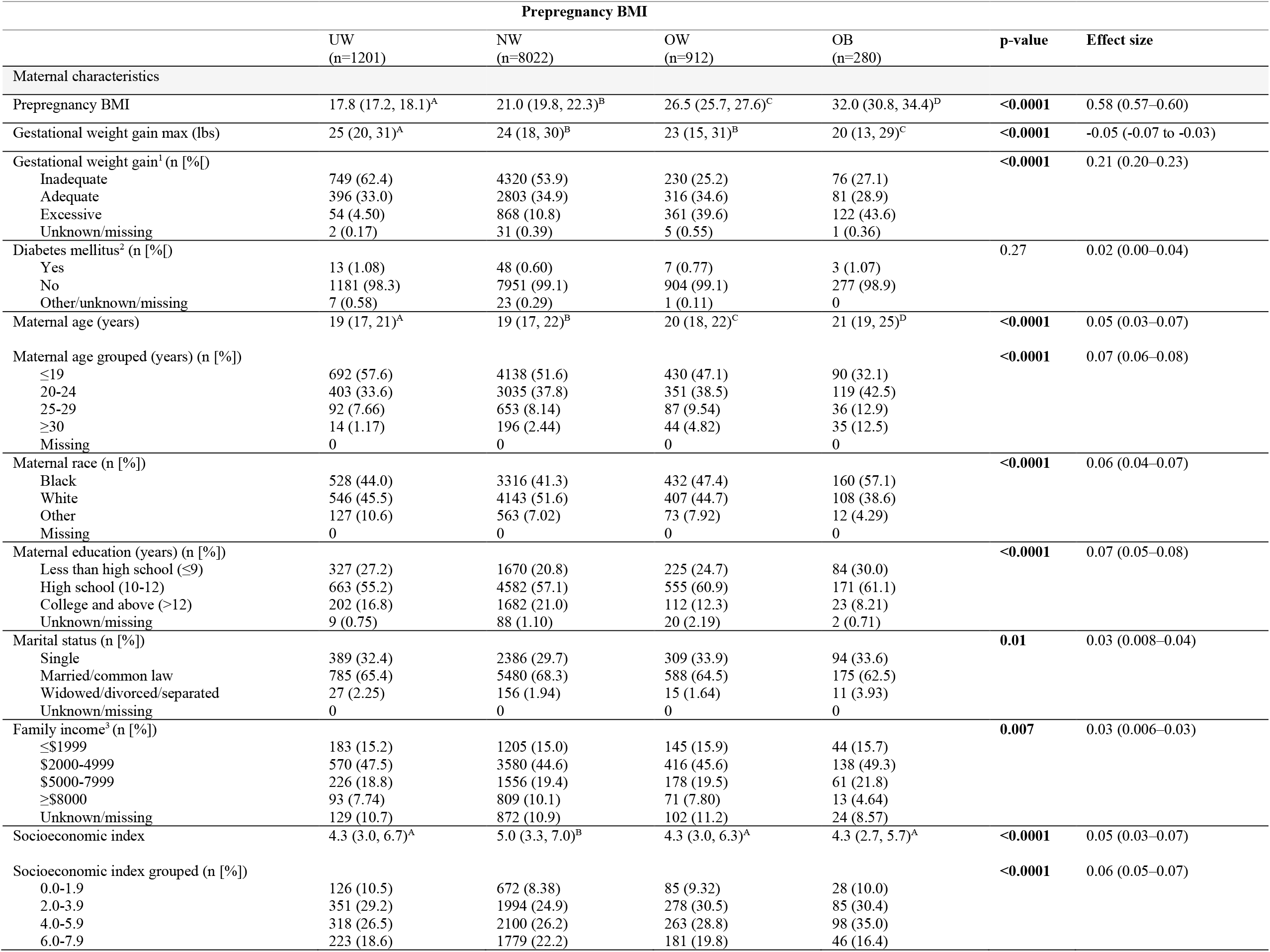
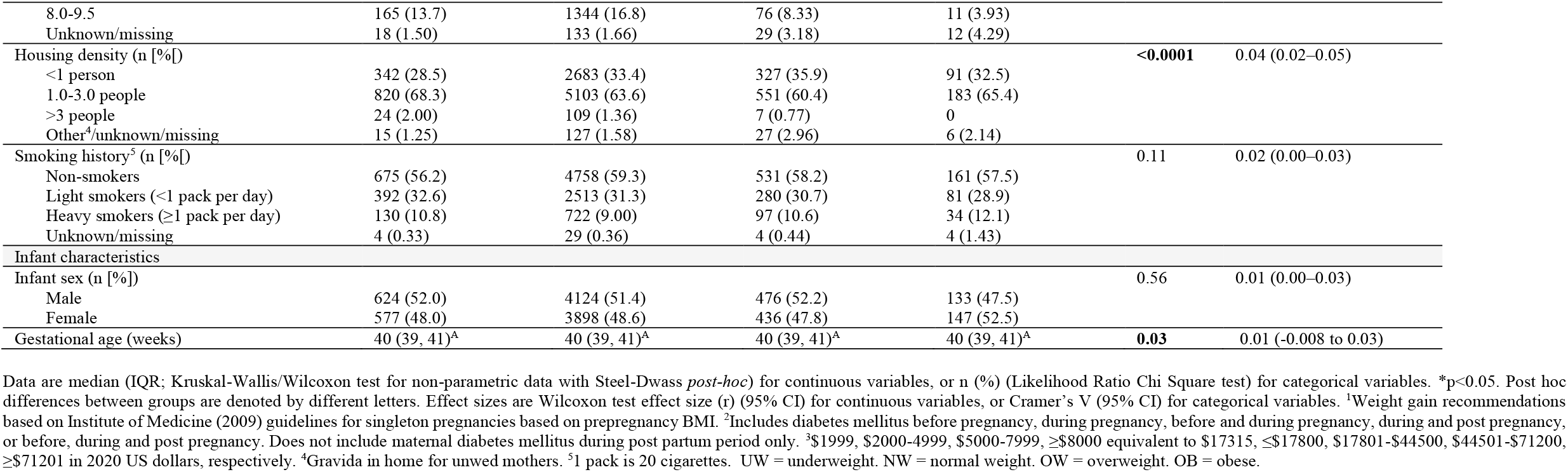
Maternal characteristics by prepregnancy BMI in term pregnancies, N=10,415.

**Supplementary Table 5.** Socioeconomic index (SES) stratified by study site.

| Study Site Location | Institution | SES Index | p-value |
| --- | --- | --- | --- |
| Boston, MA | Lying-In Hospital Children's Medical Center | 7.0 (5.3, 8.3)<sup>A</sup> | **<0.0001** |
| Buffalo, NY | Children's Hospital, SUNY | 8.3 (7.0, 9.3)<sup>B</sup> |  |
| New Orleans, LA | Charity Hospital | 3.0 (2.0, 4.0)<sup>C</sup> |  |
| New York, NY | Columbia-Presbyterian | 5.3 (4.0, 6.7)<sup>D</sup> |  |
| Baltimore, MD | Johns Hopkins Hospital | 3.3 (2.0, 5.0)<sup>C</sup> |  |
| Richmond, VA | Medical College of Virginia | 3.0 (2.0, 4.3)<sup>C</sup> |  |
| Minneapolis, MN | University of Minnesota | 7.0 (5.4, 8.3)<sup>E</sup> |  |
| New York, NY | New York Medical College | 4.0 (3.0, 5.3)<sup>F</sup> |  |
| Portland, OR | University of Oregon Medical School | 3.7 (2.7, 5.0)<sup>G</sup> |  |
| Philadelphia, PA | Pennsylvania Hospital Children's Hospital of Philadelphia | 3.7 (2.3, 5.0)<sup>CG</sup> |  |
| Providence, RI | Providence Lying-In | 4.7 (3.3, 6.3)<sup>H</sup> |  |
| Memphis, TN | University of Tennessee College of Medicine | 3.0 (2.0, 4.3)<sup>C</sup> |  |

Data are median (IQR; Kruskal-Wallis/Wilcoxon test for non-parametric data with Steel-Dwass *post-hoc*). \*p<0.05. Post hoc differences between groups are denoted by different letters.

**Supplementary Table 6.**
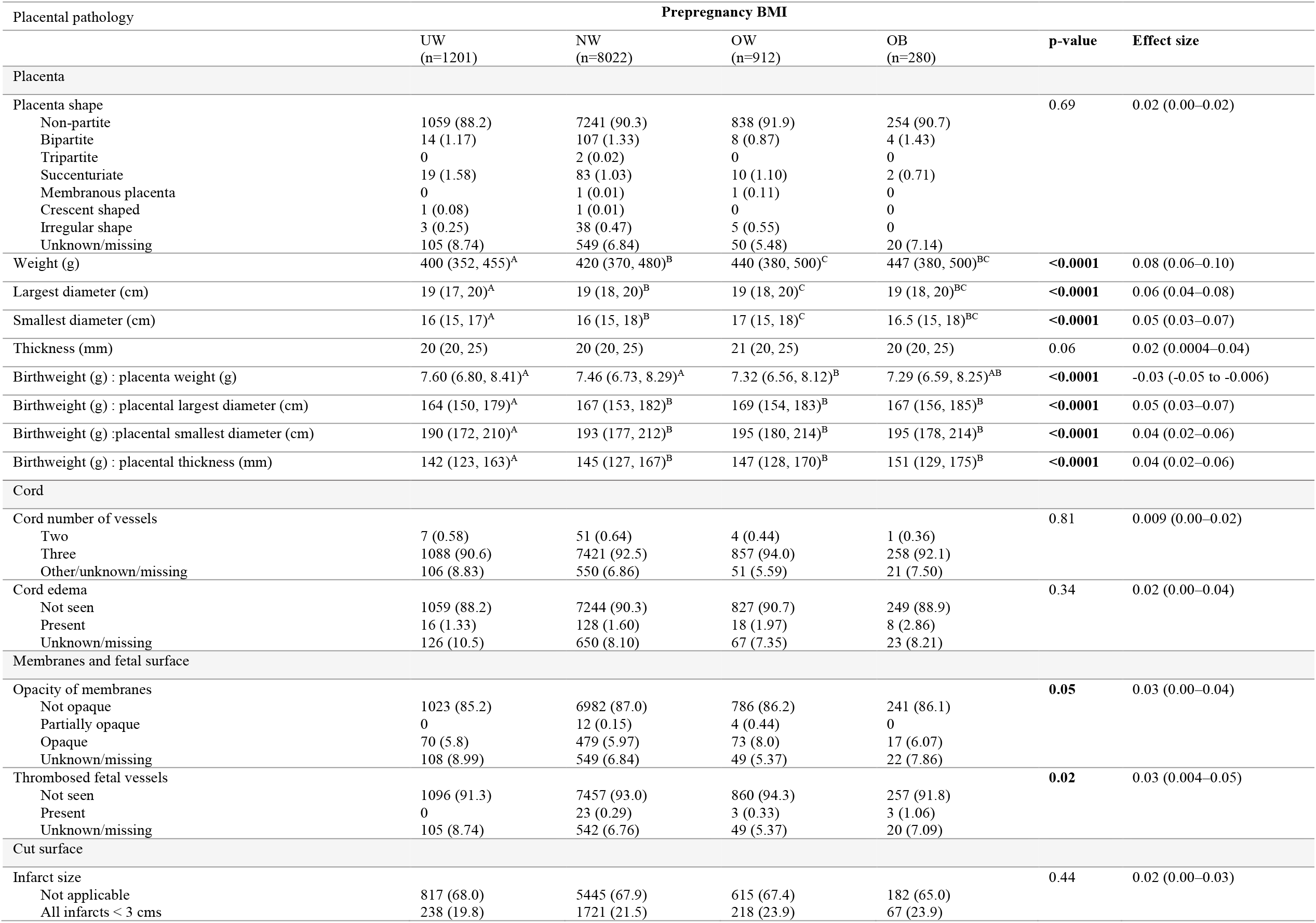
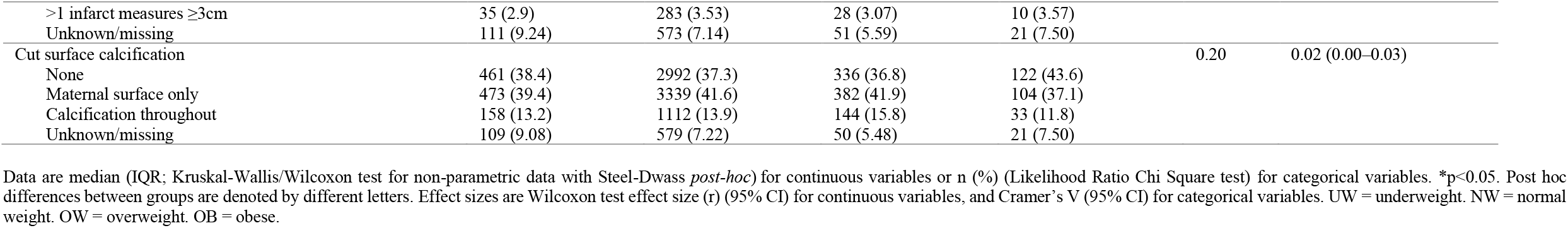
Associations between maternal prepregnancy BMI and macroscopic placental pathologies in term pregnancies, N=10,415.

**Supplementary Table 7.** Multivariable analyses for associations between maternal prepregnancy BMI (continuous) and macroscopic placental pathologies in term pregnancies, N=10,415.

| Placental pathology | Unadjusted model |  |  | Adjusted model |  |  |
| --- | --- | --- | --- | --- | --- | --- |
| | $\beta$ (95% CI) | OR (95% CI) | p-value | $\beta$ (95% CI) | OR (95% CI) | p-value |
| **Placenta** |  |  |  |  |  |  |
| Weight (g) | 2.84 (2.33–3.36) | - | **** | 3.71 (3.18–4.25) | - | **** |
| Largest diameter (cm) | 0.04 (0.03–0.06) | - | **** | 0.06 (0.04–0.07) | - | **** |
| Smallest diameter (cm) | 0.04 (0.03–0.05) | - | **** | 0.05 (0.03, 0.06) | - | **** |
| Thickness (mm) | 0.007 (-0.02 to 0.03) | - | NS | 0.05 (0.02, 0.07) | - | *** |
| Birthweight (g) : placenta weight (g) | -0.02 (-0.03 to -0.02) | - | **** | -0.02 (-0.03 to -0.02) | - | **** |
| Birthweight (g) : placental largest diameter (cm) | 0.20 (0.07–0.33) | - | ** | 0.43 (0.30–0.57) | - | **** |
| Birthweight (g) : placental smallest diameter (cm) | 0.28 (0.11–0.44) | - | *** | 0.56 (0.39–0.73) | - | **** |
| Birthweight (g) : placental thickness (mm) | 0.55 (0.35–0.76) | - | **** | 0.56 (0.34–0.77) | - | **** |
| **Cord** |  |  |  |  |  |  |
| Cord number of vessels | -0.02 (-0.10 to 0.06) | 0.98 (0.90–1.06) | NS | -0.03 (-0.12 to 0.06) | 0.97 (0.89–1.06) | NS |
| Two |  |  |  |  |  |  |
| Three |  |  |  |  |  |  |
| Cord edema | 0.03 (-0.01 to 0.07) | 1.03 (0.99–1.07) | NS | 0.02 (-0.03 to 0.07) | 1.02 (0.97–1.07) | NS |
| Not seen |  |  |  |  |  |  |
| Present |  |  |  |  |  |  |
| **Membranes and fetal surface** |  |  |  |  |  |  |
| Opacity of membranes | 0.02 (-0.002 to 0.04) | 1.02 (1.00–1.05) | NS | 0.02 (-0.008 to 0.04) | 1.02 (0.99–1.04) | NS |
| Not opaque |  |  |  |  |  |  |
| Any degree of opacity |  |  |  |  |  |  |
| Thrombosed fetal vessels | 0.06 (-0.02 to 0.15) | 1.07 (0.98–1.16) | NS | 0.05 (-0.06 to 0.16) | 1.05 (0.94–1.18) | NS |
| Not seen |  |  |  |  |  |  |
| Present |  |  |  |  |  |  |
| **Cut surface** |  |  |  |  |  |  |
| Infarcts | 0.02 (0.006–0.03) | 1.02 (1.01–1.03) | ** | 0.03 (0.02–0.04) | 1.03 (1.02–1.05) | **** |
| Not applicable |  |  |  |  |  |  |
| Any infarcts seen |  |  |  |  |  |  |
| Cut surface calcification | -0.004 (-0.02 to 0.009) | 1.00 (0.98–1.01) | NS | 0.002 (-0.01 to 0.02) | 1.00 (0.99–1.02) | NS |
| None |  |  |  |  |  |  |
| Any calcification seen |  |  |  |  |  |  |

**Supplementary Table 8.**
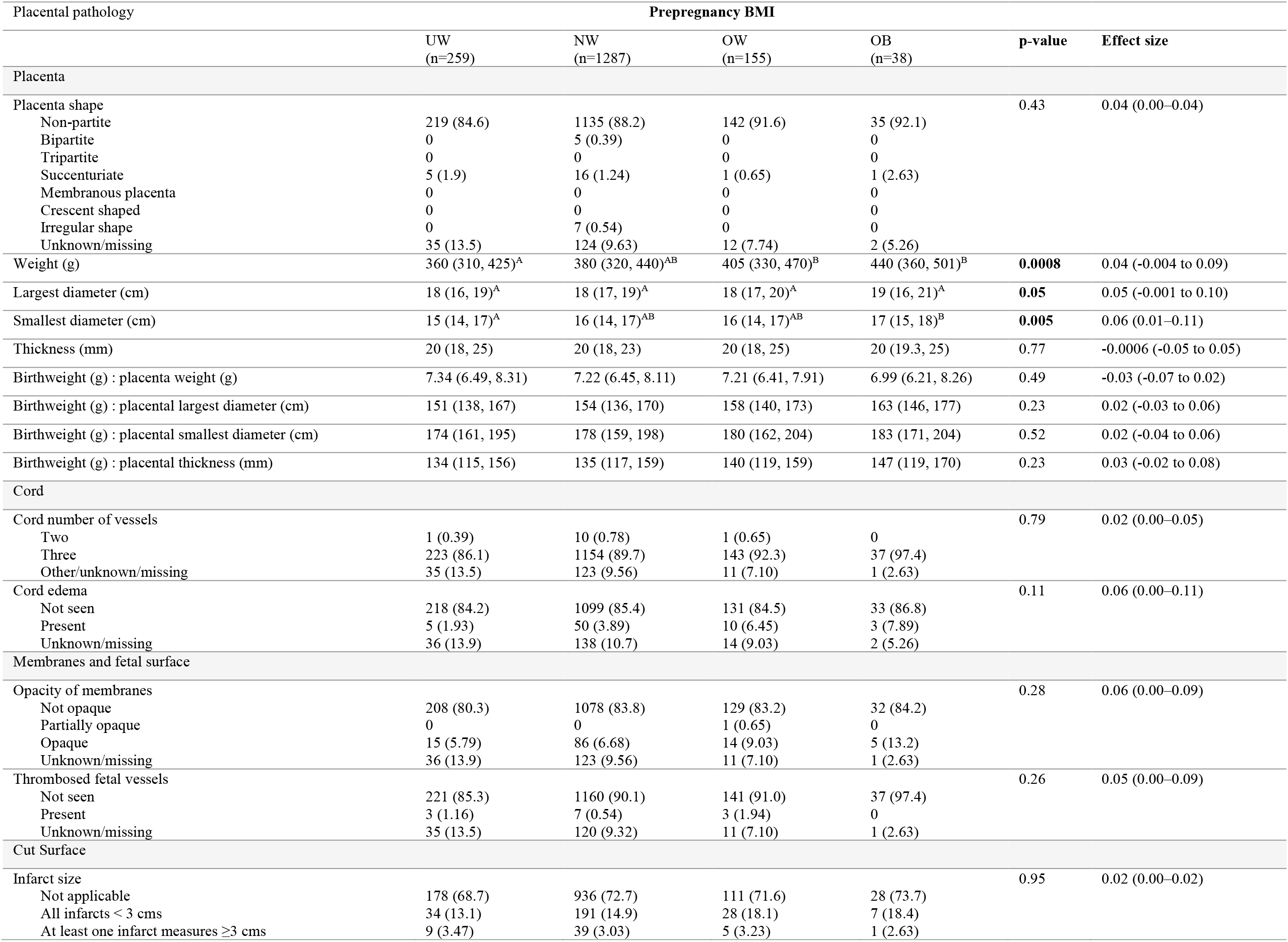
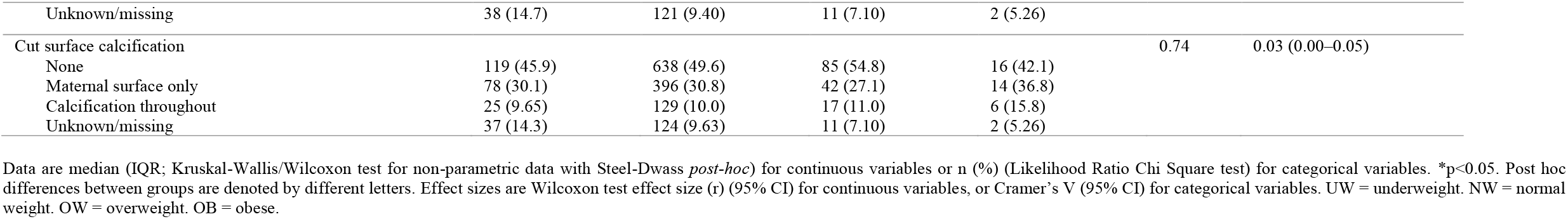
Associations between maternal prepregnancy BMI and macroscopic placental pathologies in preterm pregnancies, N=1739.

**Supplementary Table 9.**
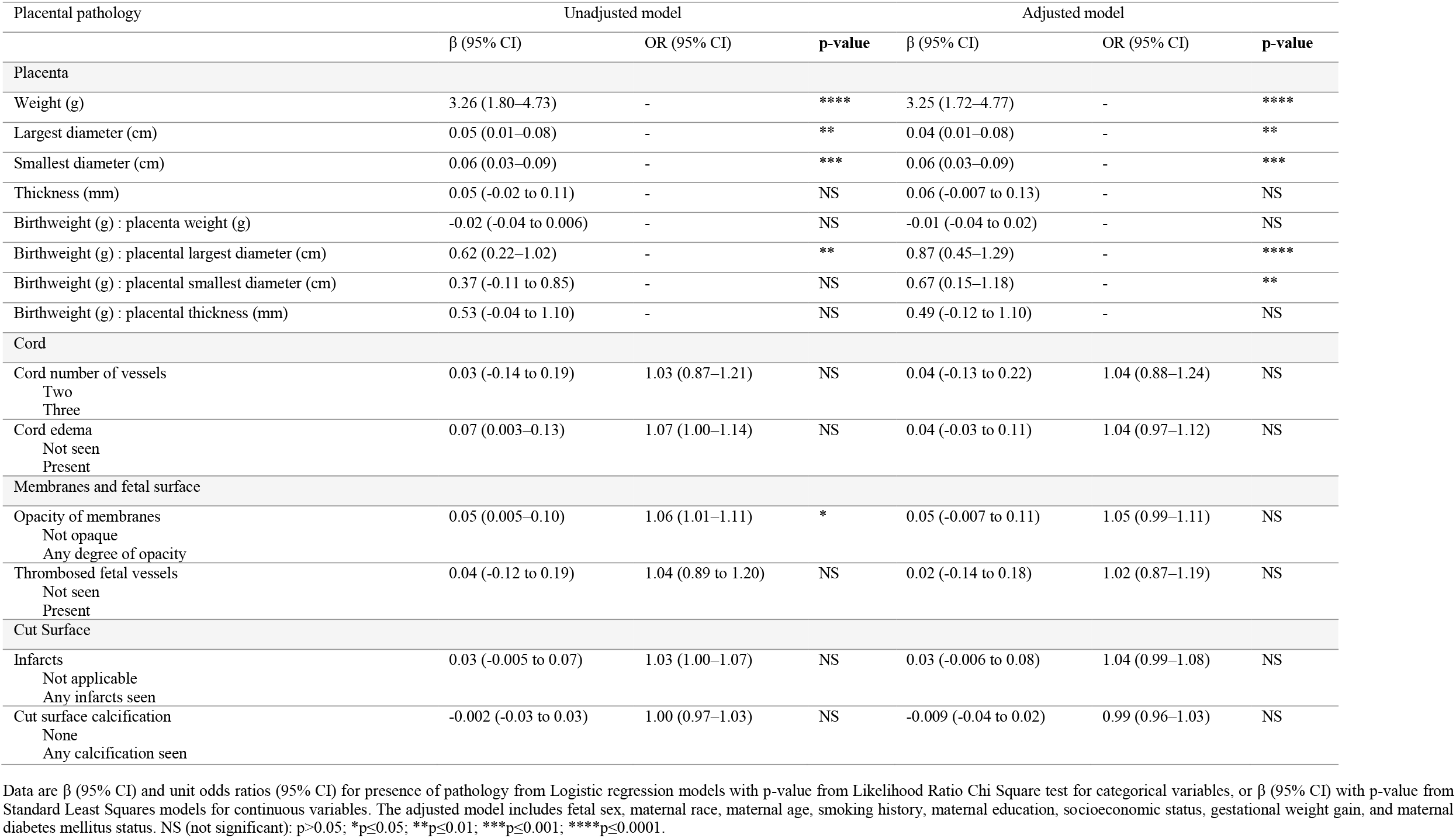
Multivariable analyses for associations between maternal prepregnancy BMI (continuous) and macroscopic placental pathologies in preterm pregnancies, N=1739.

## Supplementary figures

**Supplementary Figure 1.**
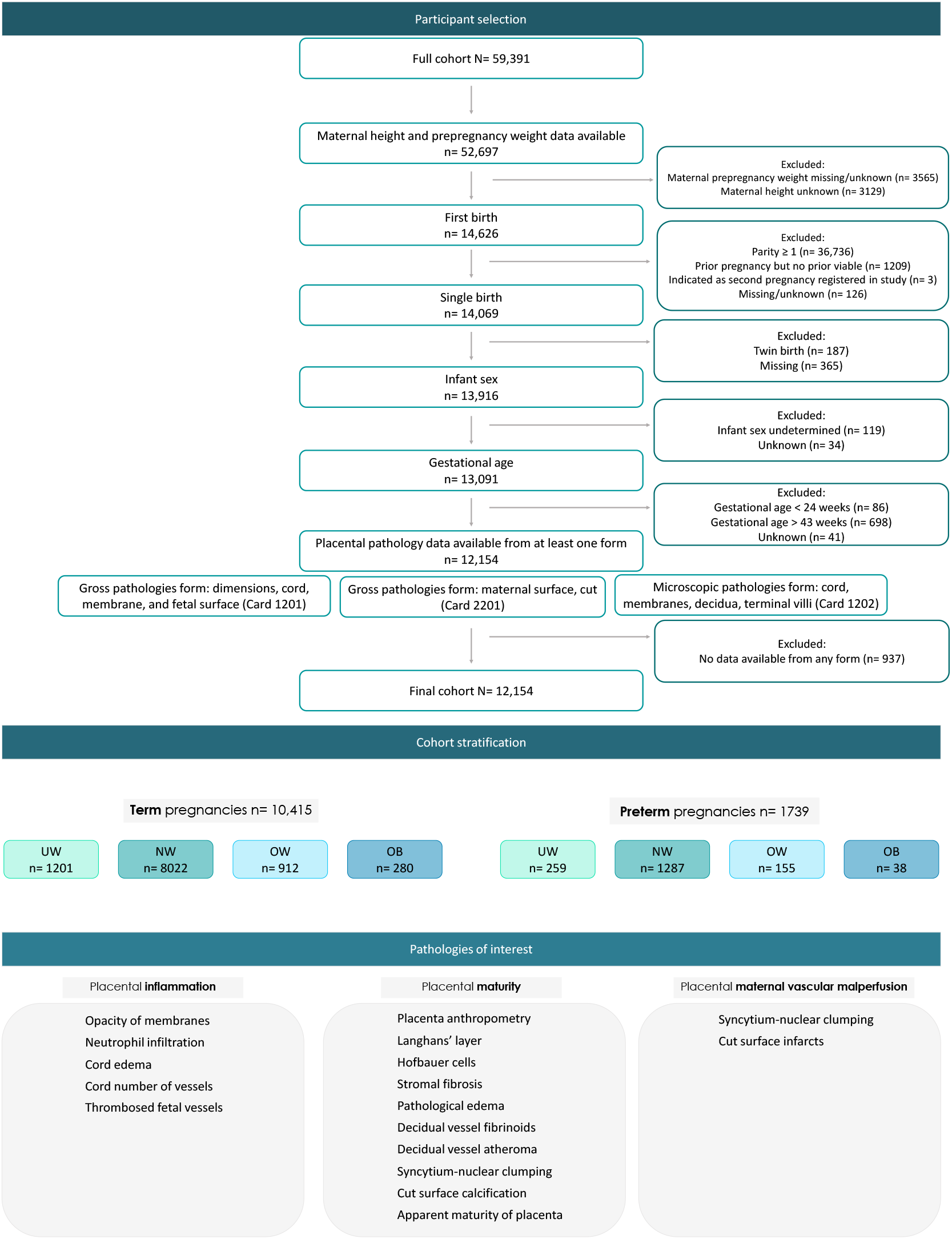
Participant flow selection and placental pathologies of interest.

**Supplementary Figure 2.**
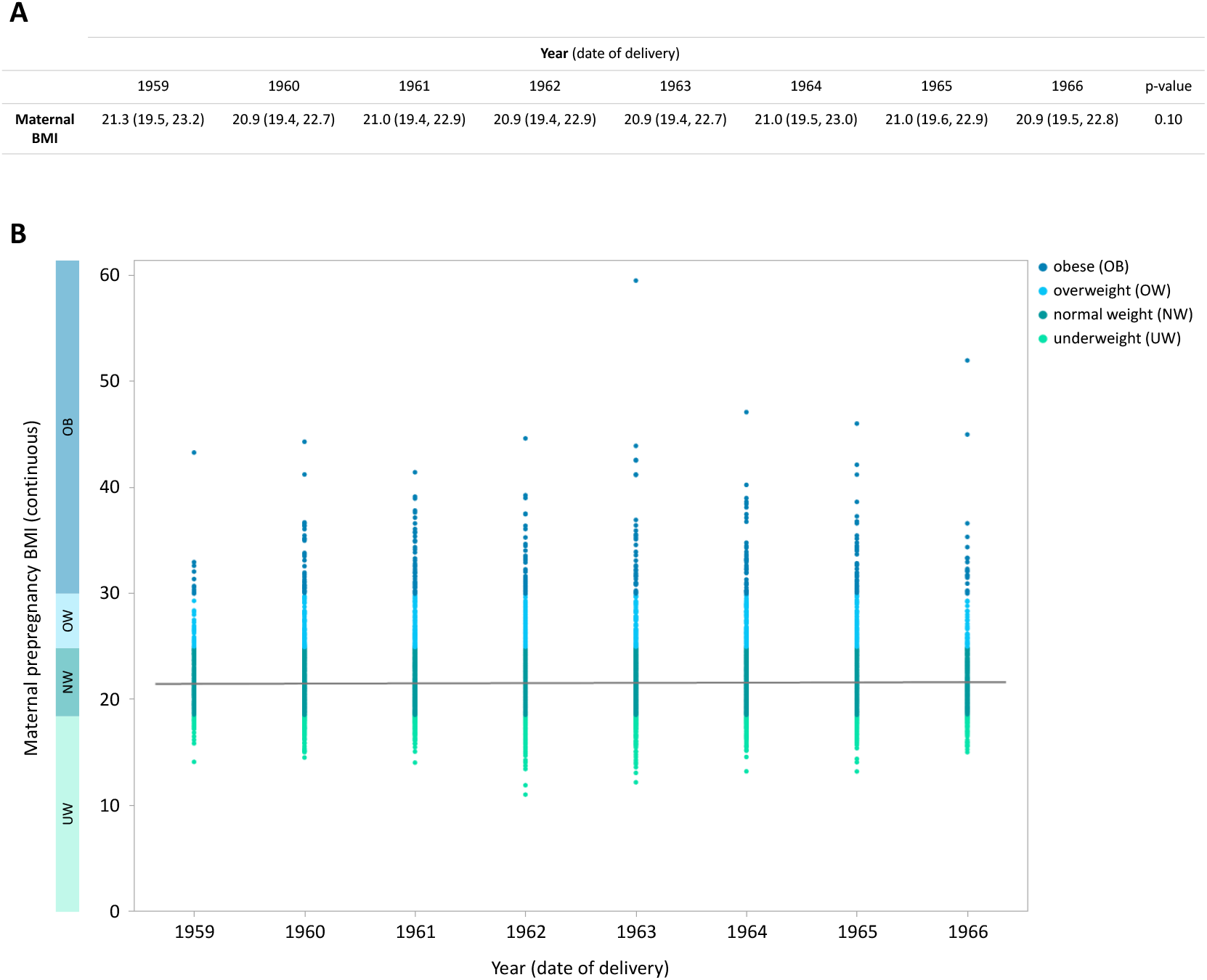
Maternal prepregnancy BMI (continuous) across study years (based on date of delivery). There were no differences in maternal BMI (continuous) stratified by year of delivery. Data are (A) median (IQR; Kruskal-Wallis/Wilcoxon test), and (B) scatter plot with line of fit.

**Supplementary Figure 3.**
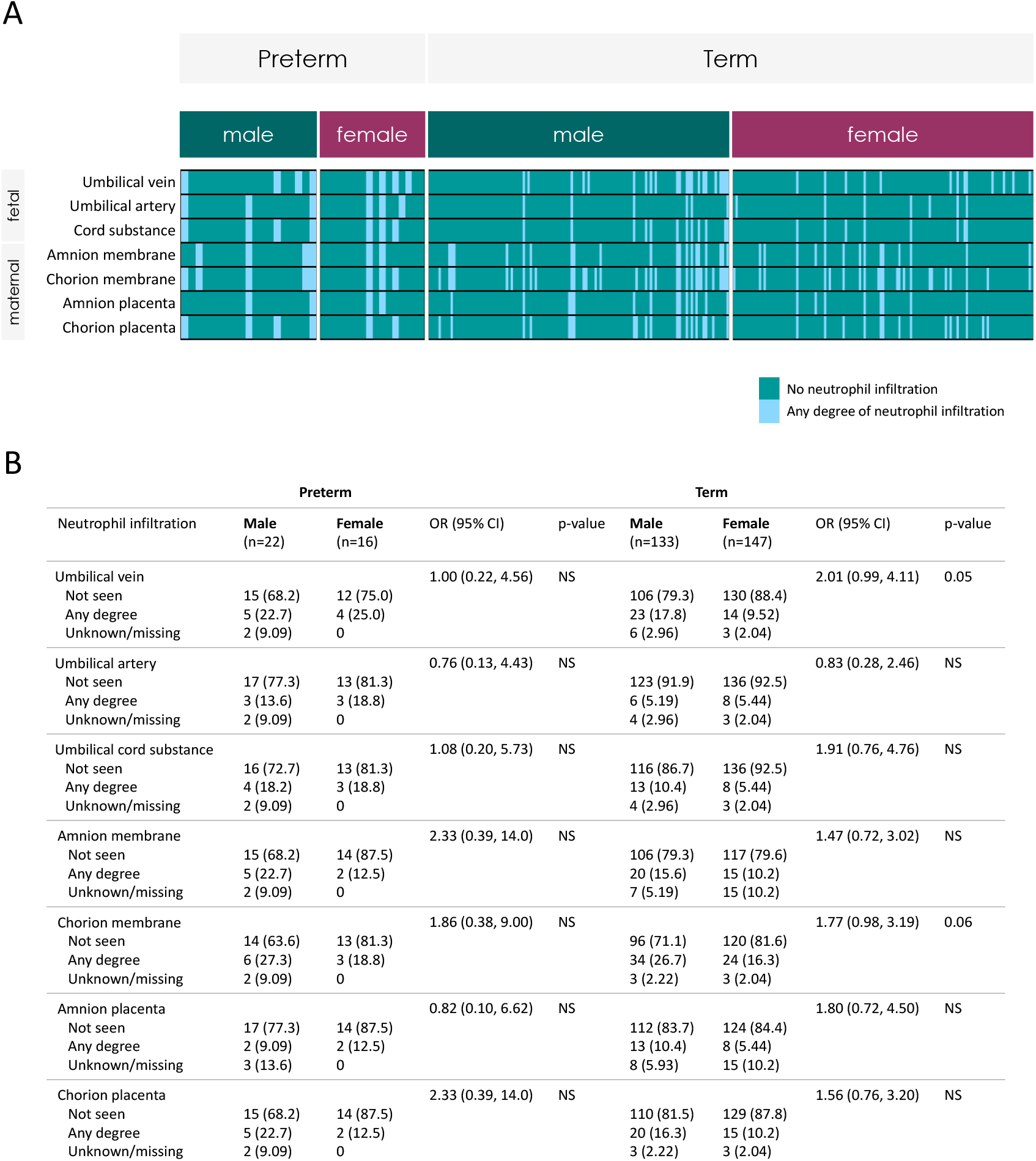
Placental neutrophil infiltration in pregnancies with maternal obesity by fetal sex, n=318. Prescence or absence of neutrophil infiltration for female and male placentae among pregnancies with maternal obesity by preterm (male: n=22, female: n=16) and term (male: n=133, female: n=147) birth. Data are n (%) (Likelihood Ratio Chi Square test) and odds ratio (95% CI) for presence of pathology in male placentae.

## Notes

### Competing Interest Statement

The authors have declared no competing interest.

### Author Declarations

Carleton University Research Ethics Board (REB) exempts publicly available and anonymous data from REB review.

